# Circulating Immune Cell Phenotypes are Associated with Socioeconomic Status and Severity of Environmental Enteropathy Among Zambian Adults

**DOI:** 10.64898/2026.08.13.26360365

**Authors:** Tracy N Phiri, Enala Musheba, Annie E Simoonga, Limpo Muyunda, Perpetual Ngalande, Mirriam Kunaka, Ian Chisenga, Mulima Mwiinga, Rose Banda, Paul Kelly, Claire D Bourke, the GI Tools Study Team

## Abstract

Environmental enteropathy (EE) is a chronic, subclinical disorder of the small intestine common in low- and middle-income countries (LMICs), where access to sanitation and exposure to enteric pathogens vary greatly by socioeconomic status (SES). Systemic immune cell activation by enteric microbial exposure is a suspected but poorly characterized driver of EE severity. We hypothesised that adults from Low-SES communities would have more severe EE than adults from High-SES communities and that this would be associated with distinct circulating immune cell phenotypes. We enrolled clinically healthy adults from High- (n=26) and Low-SES (n=76) communities in Lusaka, Zambia. Duodenal biopsies from these adults were used for microscopic morphometry assessments, while plasma and stool biomarkers of epithelial damage, intestinal inflammation, microbial translocation, and systemic inflammation were measured by ELISA. Circulating monocyte, neutrophil and T cell phenotypes were characterised in buffy coat cells by flow cytometry. Compared with the High-SES group, adults from Low-SES communities had higher duodenal villus width and crypt depth and lower epithelial surface area, indicative of more severe EE pathology, and higher levels of plasma biomarkers associated with microbial translocation and systemic inflammation. The Low-SES group also had higher expression of activation markers (CD86 and TLR4) and lower expression of HLA-DR on circulating classical monocytes and neutrophils, higher percentages of gut-homing (α4β7^+^) and activated/exhausted (PD-1^+^) T cells, including gut-homing (α4β7^+^) regulatory T cells. Principal Component Analysis identified key patterns of immune cell phenotypes across SES groups. Confounder-adjusted linear regression models showed that Principal Component 1 (monocyte/neutrophil activation) was inversely associated with duodenal villus height and epithelial surface area across SES groups. These findings indicate that EE severity varies by SES within LMIC and suggest that monocyte and neutrophil activation is linked to greater duodenal remodelling in adults with EE.

**Teaser:** Circulating innate immune cell activation phenotypes associated with duodenal features of Environmental Enteropathy are higher in adults from a Low-versus High-income community in Lusaka, Zambia, suggesting that systemic immune activation may contribute to socioeconomic disparities in gut health within LMIC.

## INTRODUCTION

Environmental enteropathy (EE) is a chronic subclinical disorder of the small intestine, ubiquitous among individuals living in areas with inadequate sanitation and persistent exposure to enteric pathogens. It is prevalent in low- and middle-income countries (LMIC), with the highest prevalence occurring in Africa, Latin America, and Asia [1–3]. In LMIC, EE severity may vary by socio-economic status (SES), with low-income communities thought to be most affected [4]. In 2022, approximately 2.2 billion people worldwide lacked access to safely managed drinking water and sanitation services; among them, 70 million lacked basic water services, while 3.5 million lacked safely managed sanitation [5]. Although EE does not present with acute symptoms, it has been linked to nutritional deficiencies [1,2,6–8], growth faltering, and reduced efficacy of oral vaccines such as rotavirus and polio [1], which contribute to broader health inequalities affecting LMICs and Low-SES communities worldwide.

Histological assessment of duodenal biopsies is the gold standard in EE diagnosis [9], which is characterised by blunted villous architecture (reduced villus height and epithelial surface area, high villus width), increased epithelial permeability and, in severe cases, breaches of the duodenal epithelial barrier [10–14]. These mucosal changes in people from Low-SES environments originate in childhood [12] and negatively impact intestinal functions, including lower nutrient absorption, and increased passage of microbes, their pathogen-associated molecular patterns (PAMPs) and other microbial products from the gut lumen into circulation, a process termed microbial translocation [15]. Adults living with EE in LMICs have intestinal dysbiosis [16], altered duodenal epithelial cell populations (e.g. fewer goblet cells and Paneth cells), infiltration of immune cells into the lamina propria and transcriptomic signatures of mucosal inflammation [1] compared to those from high-income countries, suggesting that environmentally driven changes in duodenal physiology influence gut functional capacity. However, much less is known about differences in EE and its SES-related presentation or severity within LMICs, where exposures to endemic pathogen species, seasons, and staple diets are likely to differ by SES. Several studies indicate that the circulating immune cell compartments of communities within LMIC differ according to SES and environment [17–20]. Recently, a study by Cisse *et al.* (2026) demonstrated distinct surface marker expression on myeloid cells and higher expression of cell surface markers putatively associated with senescence and inhibitory checkpoints in T cells and germinal centre formation in B cells in blood samples from adults living in Low-versus High-SES communities in Senegal [20]. However, EE is a plausible contributor and/or consequence of such SES-associated differences in circulating immune cell phenotypes which, to our knowledge, remain unexplored.

Histological evidence that immune cells accumulate in the duodenal lamina propria and the higher circulating levels of anti-microbial proteins in people with EE versus without EE suggest that affected people also have systemic immune activation. This is supported by proxy measures of inflammation and immune activation, which are elevated in blood and stool samples from people with EE [21]. For example, various studies have identified higher levels of faecal myeloperoxidase (MPO) and calprotectin, markers of neutrophil activity; plasma lipopolysaccharide (LPS)-binding protein (LBP), soluble CD14, and CD163, which are released by activated monocytes upon exposure to PAMP via their pathogen recognition receptors (PRR); and plasma endotoxin-core antibodies associated with adaptive immune responses to microbially derived LPS (endotoxin), in people with EE living in LMICs versus EE-free individuals in high-income countries [22–24]. However, most of these biomarkers are not specific to EE, as they are also elevated in other inflammatory conditions, including malaria, HIV, sepsis, obesity, rheumatoid arthritis, and inflammatory bowel disease [25–29]. Few studies have evaluated immune cells, which are the source of many of these biomarkers, in people living with EE. A small single-cell sequencing study by Kummerlowe *et al.* (2021) reported dysregulated Wingless-Related Integration Site (WNT) and Mitogen-Activated Protein Kinase (MAPK) signalling with increased proinflammatory cytokine gene expression in a tissue-resident memory T cell subset in duodenal tissue from Zambian adults with EE (n=11) versus EE-free controls from the USA (n=5) [30]. However, circulating immune cell phenotypes and their associations with duodenal morphometry remain poorly understood in EE within LMICs.

This study addresses our hypothesis that adults living in Low-SES communities have more severe EE than those from High-SES communities within the same LMIC and that this is associated with heightened activation and gut-homing phenotypes of circulating immune cells. We focus on Zambia, where more than half of the population (approximately 12.6 million people) is estimated to live below the international poverty line [31] and, as in several other LMIC [32], access to essential services, including water, sanitation, and hygiene, depends on one’s SES [33]. We enrolled adults from High- and Low-SES communities in Lusaka with the aim of answering three research questions: 1). Does duodenal morphology differ by SES? 2). Do circulating immune biomarkers and innate and adaptive immune cell phenotypes differ by SES? 3). Is there an association between circulating immune cell phenotypes and duodenal morphology? By integrating direct assessments of duodenal biopsies, systemic immune biomarkers, and innate and adaptive immunophenotyping, this study provides a comprehensive assessment of how EE-associated exposures shape circulating immune cells in adults living in High- and Low-SES settings within LMICs.

## RESULTS

### Characteristics of study participants by SES

This was a cross-sectional study that included adults from Low-SES (n=76) and High-SES (n=26) communities in Lusaka, Zambia (Table 1). Demographics did not differ significantly between groups. Significantly higher proportions of the Low-SES group were living with HIV, and lower proportions were affected by obesity (body mass index (BMI)>30) compared to the High-SES group (Table 1). Consistent with their SES grouping, a variety of wealth indicators (i.e., level of education, occupation, access to electricity, commodity ownership, and hygiene scores) differed significantly between SES groups. Clinical characteristics did not differ significantly between groups (Table S1).

**Table 1:** Characteristics of study participants from the Low- and High-SES groups.

|  |  | *High-SES*<br>*n (%)* | *Low-SES*<br>*n (%)* | *P value* |
| --- | --- | --- | --- | --- |
| *Demographics* |  |  |  |  |
|  |  | *n=26* | *n=76* |  |
| *Sex* | Male | 8 (30.8%) | 31 (41.0%) | 0.484 |
|  | Female | 18 (69.2%) | 45 (59.0%) |  |
|  |  | *n=26* | *n=76* |  |
| *Age* | 18-30 years | 14 (53.8%) | 32 (42.1%) | 0.403 |
|  | 31-40 years | 9 (34.6%) | 25 (32.9%) |  |
|  | 41-50 years | 3 (11.5%) | 12 (15.8%) |  |
|  | 51-60 years | 0 (0.0%) | 7 (9.2%) |  |
| *HIV* |  |  |  |  |
|  |  | *n=26* | *n=76* |  |
| *HIV Status* | Negative | 24 (92.0%) | 54 (71.0%) | 0.032 |
|  | Positive | 2 (8.0%) | 22 (29.0%) |  |
|  |  | *n=2* | *n=22* |  |
| *Taking ART* | Yes | 2 (100.0%) | 22 (100.0%) | 1.000 |
|  | No | 0 (0.0%) | 0 (0.0%) |  |
|  |  | *n=26* | *n=76* |  |
| *Nutritional status* |  |  |  |  |
|  | < 18.5 (Underweight) | 0 (0.0%) | 2 (2.0%) | 0.008 |
| **BMI (Kg/m<sup>2</sup>)** | 18.5-25 (Normal weight) | 9 (34.6%) | 43 (57.0%) |  |
|  | 25-30 (Overweight) | 5 (19.2%) | 21 (28.0%) |  |
|  | >30 (Obese) | 12 (46.2%) | 10 (13.0%) |  |
| **Socio-economic Factors** |  |  |  |  |
|  |  | **n=26** | **n=76** |  |
| **Level of Education** | None | 0 (0.0%) | 8 (10.5%) | <0.0001 |
|  | Primary | 1 (3.8%) | 26 (34.2%) |  |
|  | Secondary | 2 (7.7%) | 42 (55.3%) |  |
|  | College | 9 (34.6%) | 0 (0.0%) |  |
|  | University | 14 (53.8%) | 0 (0.0%) |  |
|  |  | **n=25** | **n=76** |  |
| **Occupation** | Student | 5 (20.0%) | 2 (2.6%) | <0.0001 |
|  | Formal employment | 9 (36.0%) | 11 (14.5%) |  |
|  | Business* | 8 (32.0%) | 39 (51.3%) |  |
|  | Un-employed | 3 (12.0%) | 24 (26.3%) |  |
|  |  | **n=26** | **n=76** |  |
| **House Ownership** | Yes | 12 (46.2%) | 41 (53.9%) | 0.505 |
|  | No | 14 (53.8%) | 35 (46.1%) |  |
|  |  | **n=26** | **n=76** |  |
| **Electricity** | Yes | 26 (100.0%) | 53 (69.7%) | 0.001 |
|  | No | 0 (0.0%) | 23 (30.3%) |  |
|  |  | **n=26** | **n=76** |  |
| **Car Ownership** | Yes | 21 (80.8%) | 2 (2.6%) | <0.0001 |
|  | No | 5 (19.2%) | 74 (97.4%) |  |
|  |  | **n=26** | **n=76** |  |
| **Phone Ownership** | Yes | 26 (100.0%) | 64 (84.2%) | 0.031 |
|  | No | 0 (0.0%) | 12 (15.8%) |  |
| **Lifestyle Factors** |  |  |  |  |
|  |  | **n=26** | **n=76** |  |
| **Alcohol Consumption** | Yes | 13 (50.0%) | 55 (72.4%) | 0.053 |
|  | No | 13 (50.0%) | 21 (27.6%) |  |
|  |  | **n=26** | **n=76** |  |
| **Smoking** | Yes | 4 (15.4%) | 18 (23.7%) | 0.581 |
|  | No | 22 (84.6%) | 58 (76.3%) |  |
| **Household Hygiene Factors and Practices** |  |  |  |  |
|  |  | **n=26** | **n=76** |  |
| **Boiled Water** | Yes | 3 (11.5%) | 15 (19.7%) | 0.552 |
|  | No | 23 (88.5%) | 61 (80.3%) |  |
|  |  | **n=24** | **n=75** |  |
|  | Yes | 3 (12.5%) | 29 (38.9%) | 0.023 |
| *Chlorinated Water* | No | 21 (87.5%) | 46 (61.3%) |  |
|  |  | *n=16* | *n=75* |  |
| *Hygiene Scoring\*\** | 1 | 0 (0.0%) | 0 (0.0%) | <0.0001 |
|  | 2 | 0 (0.0%) | 0 (0.0%) |  |
|  | 3 | 0 (0.0%) | 0 (0.0%) |  |
|  | 4 | 0 (0.0%) | 5 (6.7 %) |  |
|  | 5 | 0 (0.0%) | 32 (42.7%) |  |
|  | 6 | 0 (0.0%) | 14 (18.7%) |  |
|  | 7 | 0 (0.0%) | 8 (10.7%) |  |
|  | 8 | 0 (0.0%) | 6 (8.0%) |  |
|  | 9 | 2 (12.5%) | 4 (5.3%) |  |
|  | 10 | 14 (87.5%) | 0 (0.0%) |  |
| *Environmental Factors* |  |  |  |  |
| *Seasonality* | Cool/ Dry | 6 (23.1%) | 9 (11.8%) | 0.074 |
|  | Hot/ Dry | 4 (15.4%) | 29 (38.2%) |  |
|  | Wet/ Rainy | 16 (61.5%) | 38 (50.0%) |  |

Categorical variables are expressed as percentages.
All categorical variables were compared between SES groups by Fisher's exact with P values < 0.05 considered significant
ART: Antiretroviral therapy
Seasons: Rainy; December- April, Cool/ Dry; May- August, and Hot/ Dry; September to November
\*Business: Refers to informal sector traders, or owner operated.
\*\*Hygiene score; A composite of general household hygiene (2), sanitation practices (2), water storage (2), food storage (2), and hand-washing practices (2); where 1–2= Poor; 3–4= Below acceptable; 5= Acceptable / Average; 6–7= Good; 8–9= Excellent and 10= Perfect

### EE is more severe in people living in Low-SES versus High-SES communities

We hypothesised that duodenal features of EE would be more severe in the Low-SES group than the High-SES group. Using H&E-stained duodenal sections, we found that villus height did not differ significantly, while villus width (adjusted β coefficient (adj β) = 0.3, 95% confidence interval (CI) 0.2, 0.4; p<0.001) and crypt depth (adj β = 0.2, 95% CI 0.1, 0.3; p=0.005) were higher in the Low-than the High-SES group. Epithelial surface area was lower in the Low-than the High-SES group (adj β = -0.3, 95% CI -0.4, -0.1; p=0.002) after adjusting for confounders (age, sex, BMI, seasonality, alcohol consumption, and HIV status) (Fig. 1A-F and Table S2). Plasma concentrations of intestinal fatty acid binding protein (I-FABP) are used as an indirect indicator of epithelial damage, and we found some evidence that these were higher in the Low-versus High-SES group in unadjusted analyses (adj β= 0.3: 95% CI 0.1, 0.6; p= 0.012). However, I-FABP did not differ significantly (adj β= 0.2: 95% CI 0.0, 0.5; p= 0.112) between SES groups after adjusting for confounders. Faecal MPO levels did not differ significantly between the two groups in both unadjusted and adjusted analyses (Fig. 1G-H and Table S2).

**Fig. 1.**
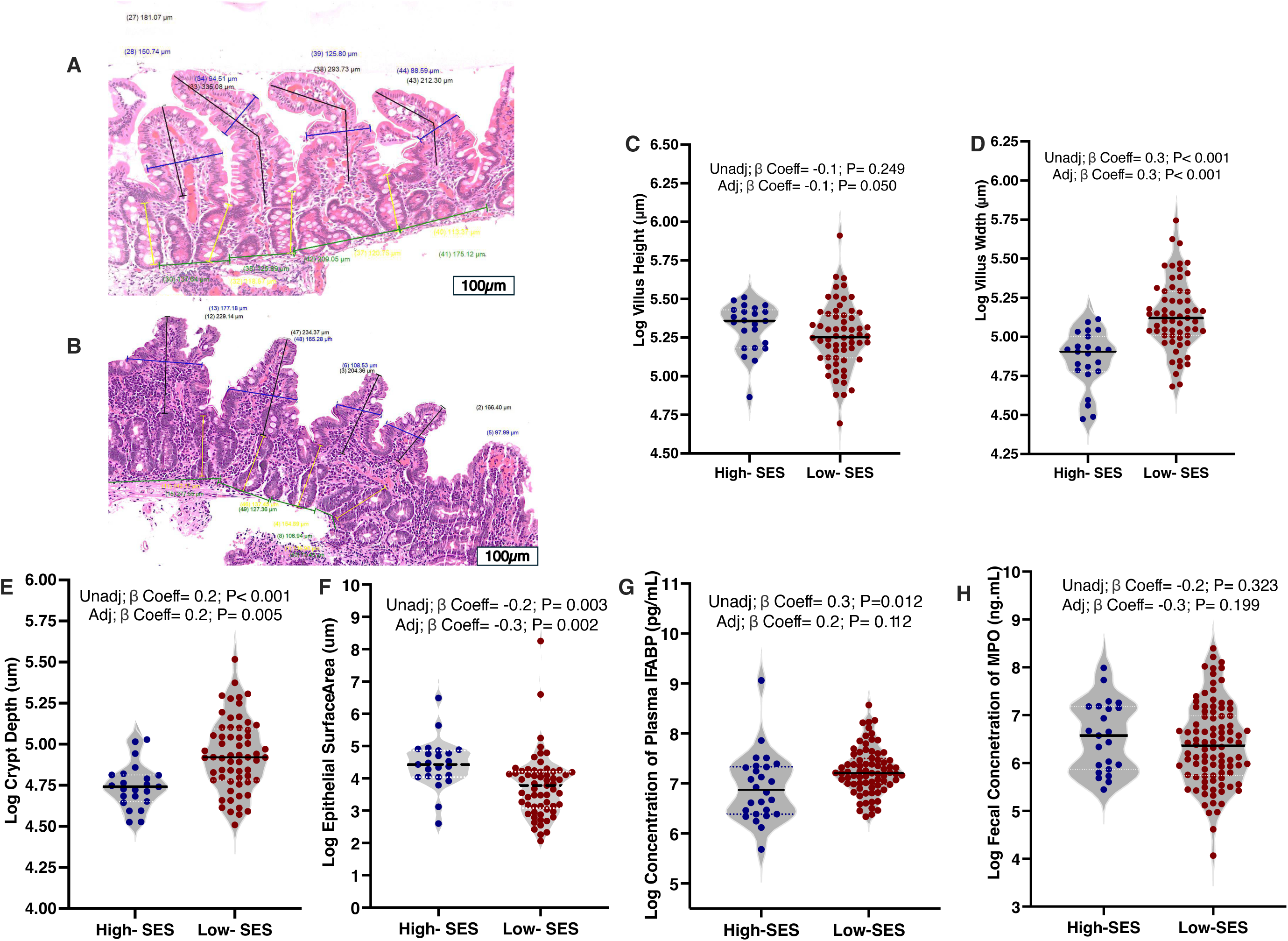
EE severity differs by SES group. A. and B. H&E staining of duodenal biopsies from adults from the High- and Low-SES groups, respectively, showing duodenal morphometry measurements (x20 magnification); Blue: Villus width, Black: Villus height, White: Epithelial surface area, and Yellow: Crypt depth. Duodenal morphometry measurements are compared between the High-(n=23) and Low-(n=61) SES groups for C. villus height, D. villus width, E. crypt depth, and F. epithelial surface area. G. Plasma concentration of I-FABP (High-SES: n= 25 and Low-SES: n= 75). H. Faecal concentrations of MPO (High-SES: n= 23 and Low-SES: n= 76). Associations between indicators of EE severity and SES group were evaluated using unadjusted and adjusted (for age, sex, BMI, seasonality, alcohol consumption, and HIV status) linear regression models, with the High-SES group as the reference. Benjamini-Hochberg (BH) adjusted P values are shown, with values < 0.05 considered significant.

### Systemic inflammation and microbial translocation are higher in adults from a Low-SES versus those from a High-SES community

We hypothesised that adults from Low-SES would have higher levels of systemic inflammation and microbial translocation than those from High-SES. Using ELISA to measure plasma levels of C-reactive protein (CRP) and alpha-1-glycoprotein (AGP), we found that CRP (adj β= 0.9: 95% CI 0.2, 1.6; p= 0.022) was higher in adults from the Low-SES than those from the High-SES group, while AGP concentrations did not differ significantly between the two groups (Fig 2A and Table S2). We went on to quantify plasma concentrations of biomarkers associated with innate immune activation in response to microbial translocation (sCD14, sCD163, and LBP) and antibodies directed at bacterial endotoxin (EndoCAbs IgG, IgA, and IgM). As hypothesised, we found significantly higher plasma concentrations of sCD14 (adj β= 1.5: 95% CI 1.3, 1.7; p< 0.001) and LBP (adj β= 0.7: 95% CI 0.4, 1.0; p< 0.001) in the Low-SES group, while sCD163 (adj β= 0.3: 95% CI 0.0, 0.5; p= 0.056) did not differ significantly between the two groups after adjustment (Fig. 2B and Table S2). EndoCabs IgG (adj β= 2.2: 95% CI 1.7, 2.6; p< 0.001) and IgM (adj β= 0.5: 95% CI 0.2, 0.9; p= 0.015) were higher in the Low-SES group, while IgA (adj β= 0.4: 95% CI 0.0, 0.8; p= 0.053) did not differ between the two groups after adjustment (Fig. 2C and Table S2).

**Fig 2:**
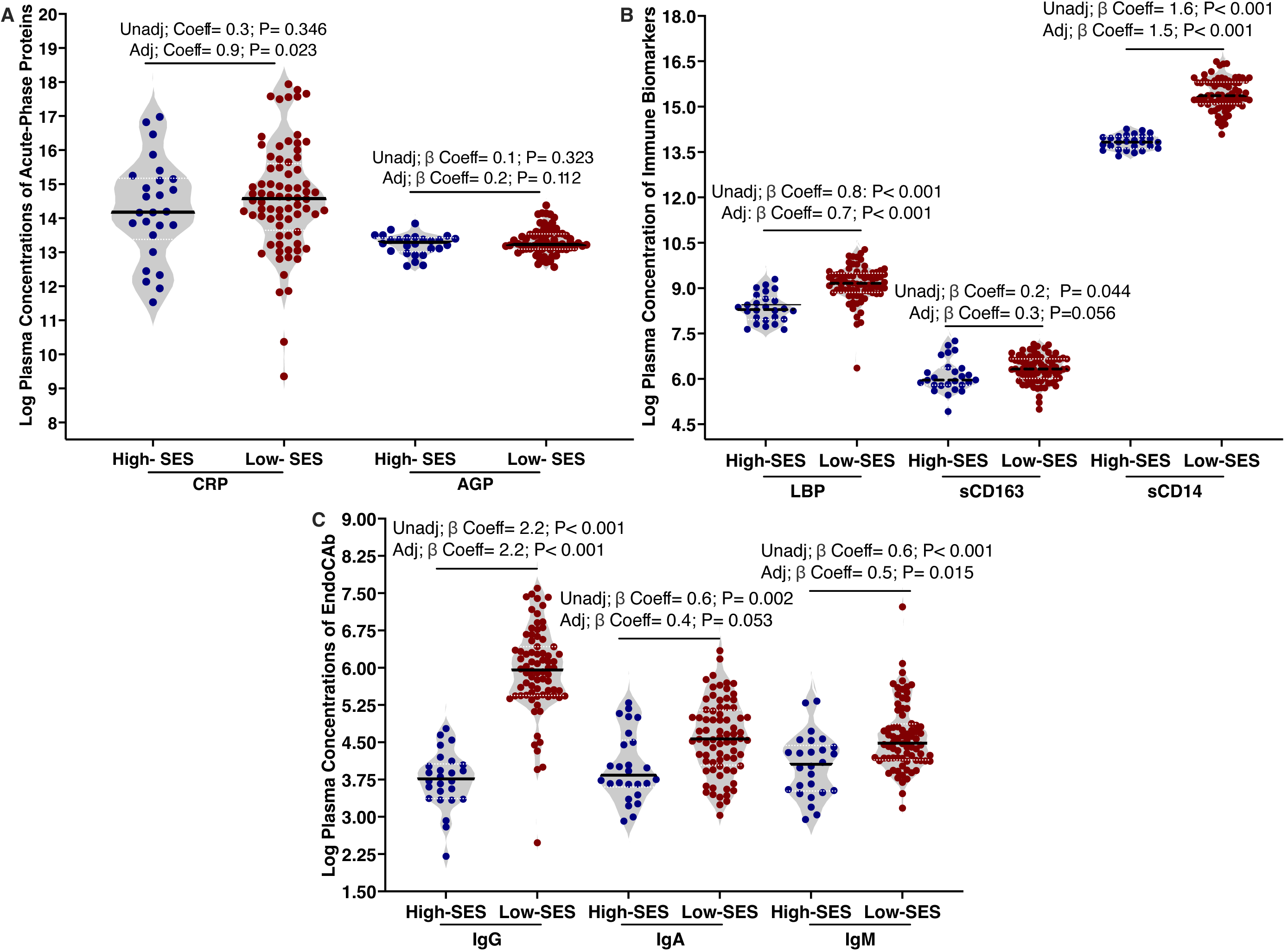
Adults with EE from the Low-SES group have higher concentrations of microbial translocation and systemic inflammation biomarkers than those from the High-SES group. A. Plasma concentrations of CRP (left) and AGP (right). B. Plasma concentrations of LBP (left), sCD163 (centre), and sCD14 (right). C. Plasma concentrations of EndoCabs IgG (left), IgA (centre), and IgM right) in the High- and Low-SES groups. Associations between biomarkers and SES group were evaluated using unadjusted and adjusted (for age, sex, BMI, seasonality, alcohol consumption, and HIV status) linear regression models, with the High-SES group as the reference. GMU (IgG Median Units); AMU (IgA Median Units); MMU (IgM Median Units); CRP, AGP and sCD14 (pg/ml); LBP and sCD163 (ng/ml); BH-adjusted P values are shown, with values < 0.05 considered significant. High-SES: (n=26) and Low-SES (n=76).

### Monocytes and neutrophils from adults with EE from Low-SES environments have a more activated phenotype

Having identified that plasma levels of the soluble monocyte-derived receptor, sCD14, and the LPS-binding protein, LBP, were higher in the Low-SES group, we hypothesised that blood monocytes from these adults would also have a more activated phenotype than those from the High-SES group. Using flow cytometry of buffy coat cell samples from both groups, we quantified monocyte subsets (Fig. 3A) and their surface expression of the LPS PRR, Toll-like receptor (TLR)-4, the major histocompatibility antigen HLA-DR, and the co-receptor CD86, which are required for antigen presentation and co-stimulation of T cells, respectively. Percentages of total monocytes (Lymphocyte lineage marker (Lin)^-^CD66b^-^HLA-DR^+^) and monocyte subclasses; classical (CD14^high^CD16^-^), intermediate (CD14^high^CD16^+^), and non-classical (CD14^low^CD16^+^) monocytes did not differ significantly between the two groups (Fig. 3B, 3C and Table S2). Total monocytes from adults in the Low-SES group had higher median fluorescence intensity (MFI) of CD86 (adj β= 0.5: 95% CI 0.4, 0.6; p< 0.001) and TLR4 (adj β= 1.0: 95% CI 0.4, 1.5; p< 0.001) but lower HLA-DR (adj β= -0.8: 95% CI -1.4, -0.3; p=0.007) than the High-SES group (Fig. S4 and Table S2). Within the monocyte subsets, CD86 expression on classical (adj β= 0.5: 95% CI 0.4, 0.6; p< 0.001), intermediate (adj β= 0.5: 95% CI 0.4, 0.6; p< 0.001), and non-classical (adj β= 0.4: 95% CI 0.3, 0.5; p< 0.001) monocytes was also higher in the Low-SES versus High-SES group. TLR-4 expression on the classical (adj β= 0.3: 95% CI 0.2, 0.3; p< 0.001), intermediate (adj β= 0.4: 95% CI 0.3, 0.5; p< 0.001), and non-classical monocytes (adj β= 0.5: 95% CI 0.4, 0.7; p< 0.001) was also higher in the Low-SES than the High-SES. HLA-DR expression on classical (adj β= -0.2: 95% CI -0.4, - 0.1; p= 0.011) was lower in the Low-versus High-SES group, while expression on intermediate and non-classical subsets did not differ significantly between the two groups (Fig 3D-F and Table S4).

**Fig 3:**
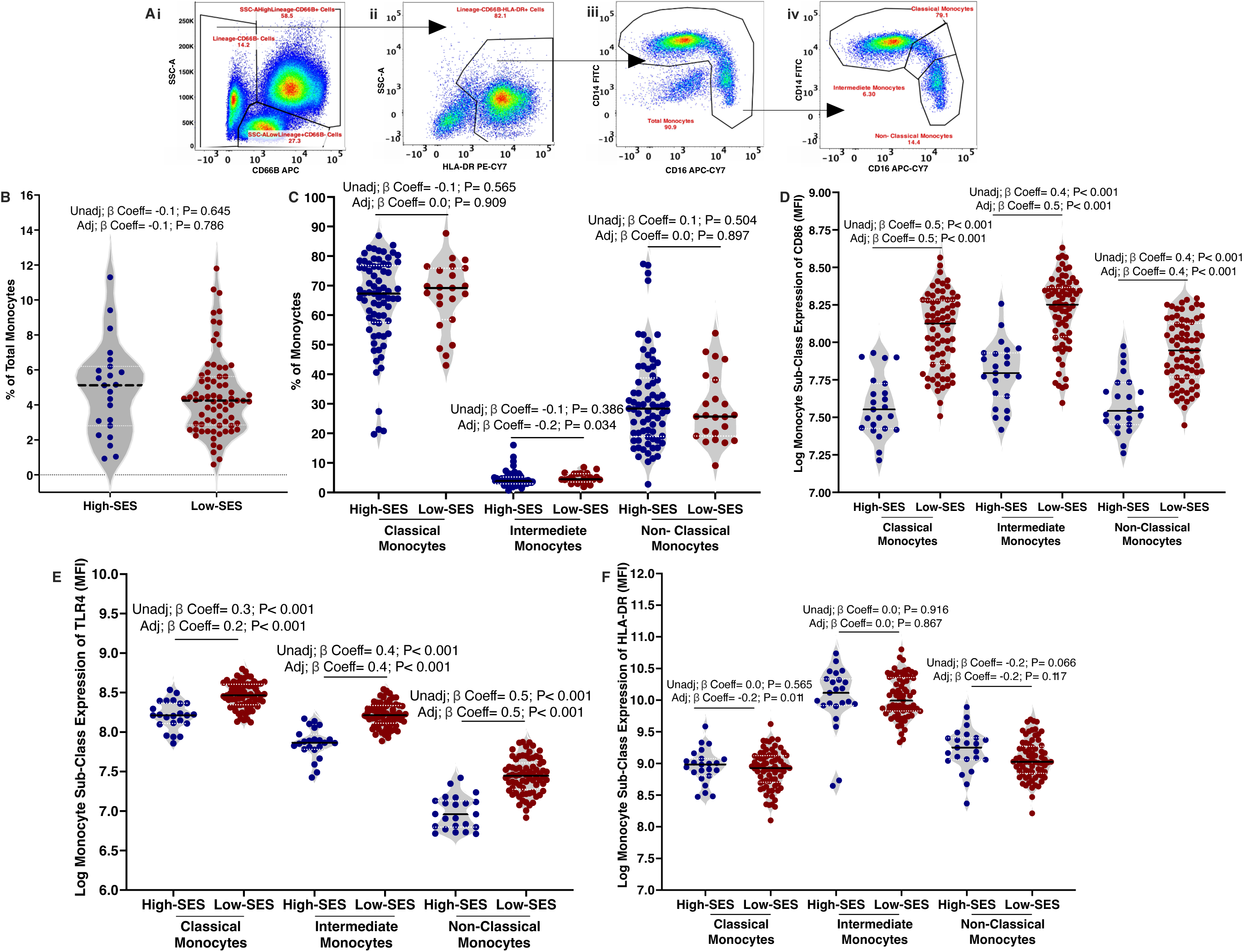
Monocytes from adults from a Low-SES community have a more activated phenotype than those from a High-SES community. A. Monocyte gating strategy: showing steps used in the identification of monocytes in a single example(left-to-right): i. Side scatter-Area (SSC-A) versus CD66B/ Lineage-APC. ii. SSC-A versus HLA-DR. iii. Total monocyte gate (CD14-FITC versus CD16-APC-CY7). iv. Monocyte subclasses (CD14-FITC versus CD16-APC-CY7). Each gate is presented as a percentage, while MFI were used to evaluate CD86, TLR-4, and HLA-DR expression on neutrophils. B. Percentages of total monocytes (Lineage^-^CD66B^-^HLA-DR^+^). C. Percentages of monocyte subclasses (left-to-right: classical (CD14^high^CD16^-^), intermediate (CD14^high^CD16^+^), and non-classical (CD14^low^CD16^+^) monocytes). D. Monocyte sub-class expression of CD86 (median fluorescence intensity, MFI). (Left-to-right): Classical, intermediate and non-classical monocytes. E. Monocyte sub-class expression (MFI) of TLR-4 (Left-to-right): Classical, intermediate and non-classical monocytes. and F. Monocyte sub-class expression (MFI) of HLA-DR (left-to-right): Classical, intermediate, and non-classical monocytes in the High- and Low-SES groups. Associations between monocyte phenotypes and SES group were evaluated using unadjusted and adjusted (for age, sex, BMI, seasonality, alcohol consumption, and HIV status) linear (MFI) and fractional (percentages) regression models, with the High-SES group as the reference. BH-adjusted P values are shown, with values < 0.05 considered significant. High-SES: (n=23) and Low-SES: (n=73).

Typically, neutrophils express very low levels of CD86 and HLA-DR in healthy individuals; however, these molecules are upregulated during infection and inflammation. Given that EE is an inflammatory condition and that neutrophils rapidly respond to PAMP and infiltrate tissue sites of inflammation, we hypothesised that neutrophils would also exhibit a more activated phenotype in the Low-versus High-SES group. As with monocytes, the percentages of total neutrophils (CD66b^+^CD16^+^; Fig. 4A) did not differ significantly between the two groups (Fig. 4B). Neutrophil expression of CD86 (adj β= 0.2: 95% CI 0.1, 0.3; p= 0.002) and TLR-4 (adj β= 0.5: 95% CI 0.4, 0.6; p< 0.001) was higher in the Low-SES versus the High-SES. HLA-DR was lower on neutrophils from the Low-SES group, compared to the High-SES group (adj β=-0.4: 95% CI -0.5, -0.2; p< 0.001); (Fig. 4C and supplementary Table 6).

**Fig 4:**
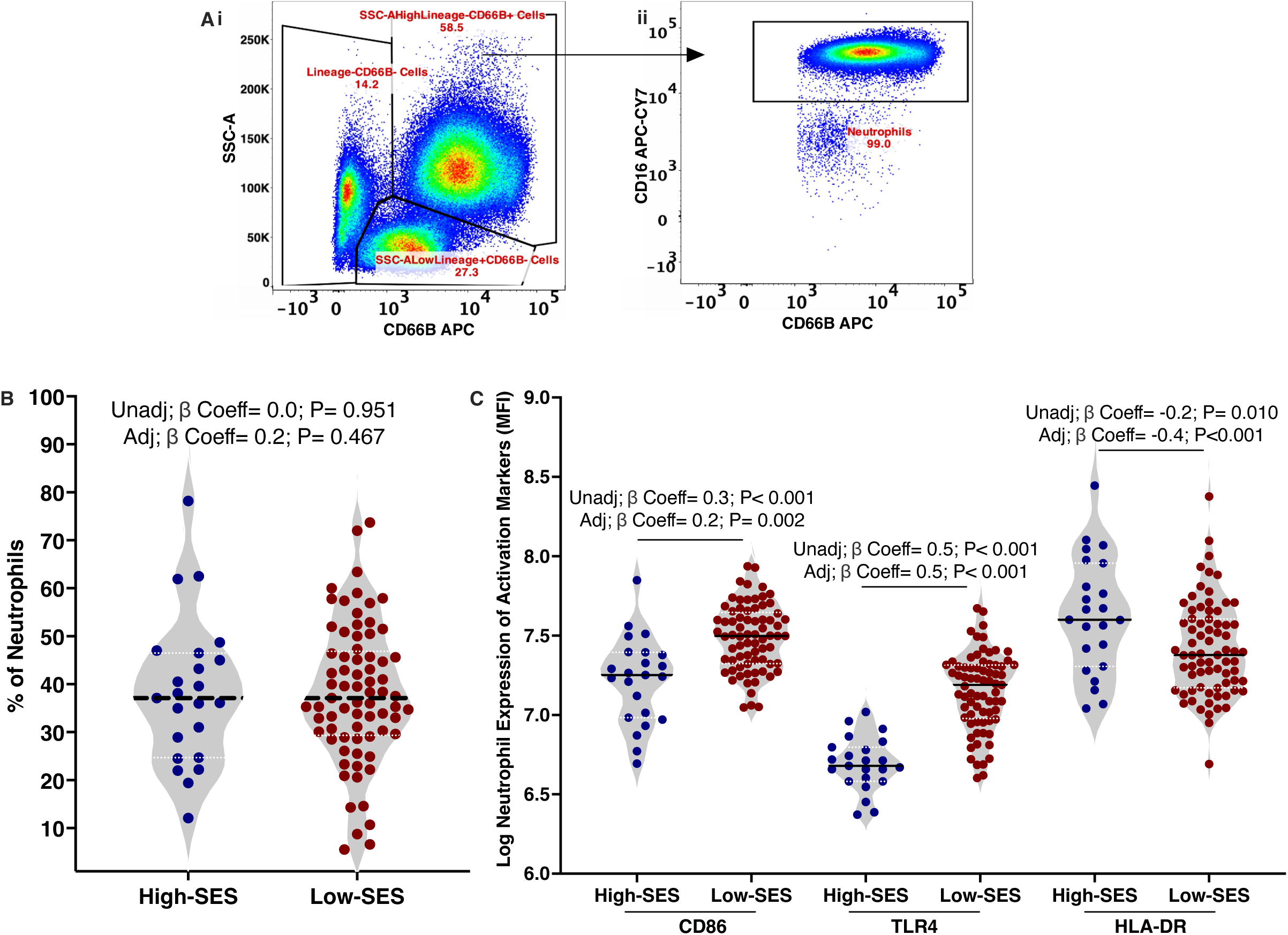
Neutrophil expression of CD86 and TLR4 is higher in adults from Low-versus High-SES communities. A. Neutrophil (SSC-A^high^CD66B+CD16^+^) flow cytometry gating strategy. Showing steps used in the identification of neutrophil cells as percentages (left-to-right) i. SSC-A versus CD66B/ Lineage-APC. ii. Total neutrophil gate (CD16-APC-CY7 vs CD66B). Each gate is presented as a percentage, while MFI were used to evaluate CD86, TLR-4, and HLA-DR expression on neutrophils. B. Percentage of neutrophils. C. Neutrophil expression (MFI) of CD86, TLR4, and HLA-DR (left-to-right) in the High- and Low-SES groups. Associations between neutrophil phenotypes and SES group were evaluated using both unadjusted and adjusted (for age, sex, BMI, seasonality, alcohol consumption, and HIV status) linear (MFI) and fractional (percentages) regression models, with the High-SES group as the reference. BH-adjusted P values are shown, with values < 0.05 considered significant. High-SES: (n=23) and Low-SES: (n=73).

### Adults with EE from the Low-SES group have higher percentages of gut-homing and exhausted CD4^+^ and CD8^+^ T cells

Having shown that people living with EE in Low-SES environments have elevated systemic inflammation but lower expression of the key antigen-presenting molecule HLA-DR on classical monocytes and neutrophils, we hypothesised that T cell phenotypes would also differ between the Low-versus High-SES group. We quantified expression of activation/exhaustion marker (programmed cell death protein-1; PD-1^+^) and integrins associated with homing to the small intestine (α4β7^+^) on CD4^+^ (CD3^+^CD4^+^) and CD8+ (CD3^+^CD4^-^) T cells, and α4β7^+^ expression on CD4^+^ T regulatory cells (Treg; CD127^-/orlow^FOXP3^+^) using flow cytometry (Fig. 5A). Percentages of total CD4+ and CD8+ T cells did not differ significantly between the two groups (Fig. 5B and Table S2). Percentages of total CD4+ T cells with a gut-homing (adj β= 0.8: 95% CI 0.4, 1.1; p< 0.001), activated/exhausted (adj β= 1.1: 95% CI 0.7, 1.5; p< 0.001), or co-expressing both the gut-homing and activated/exhausted phenotype (adj β= 0.6: 95% CI 0.2, 1.1; p= 0.012) were higher in the Low-SES group than in the High-SES group (Fig. 5C and Table S2). We also found that percentages of total Tregs (adj β= 1.1: 95% CI 0.6, 1.5; p< 0.001) and gut-homing Tregs (adj β= 1.0: 95% CI 0.4, 1.6; p= 0.004) were higher in the Low-than High-SES group (Fig. 5D and Table S2). Percentages of total CD8^+^ T cells with a gut-homing (adj β= 1.1: 95% CI 0.6, 1.6; p< 0.001), activated/ exhausted (adj β= 1.3: 95% CI 0.9, 1.7; p< 0.001), or expressing markers of both the gut-homing and exhausted/ activated phenotype (adj β= 0.6: 95% CI 0.1, 1.2; p= 0.044) were higher in the Low-SES group than the High-SES group (Fig. 5E and Table S6).

**Fig 5:**
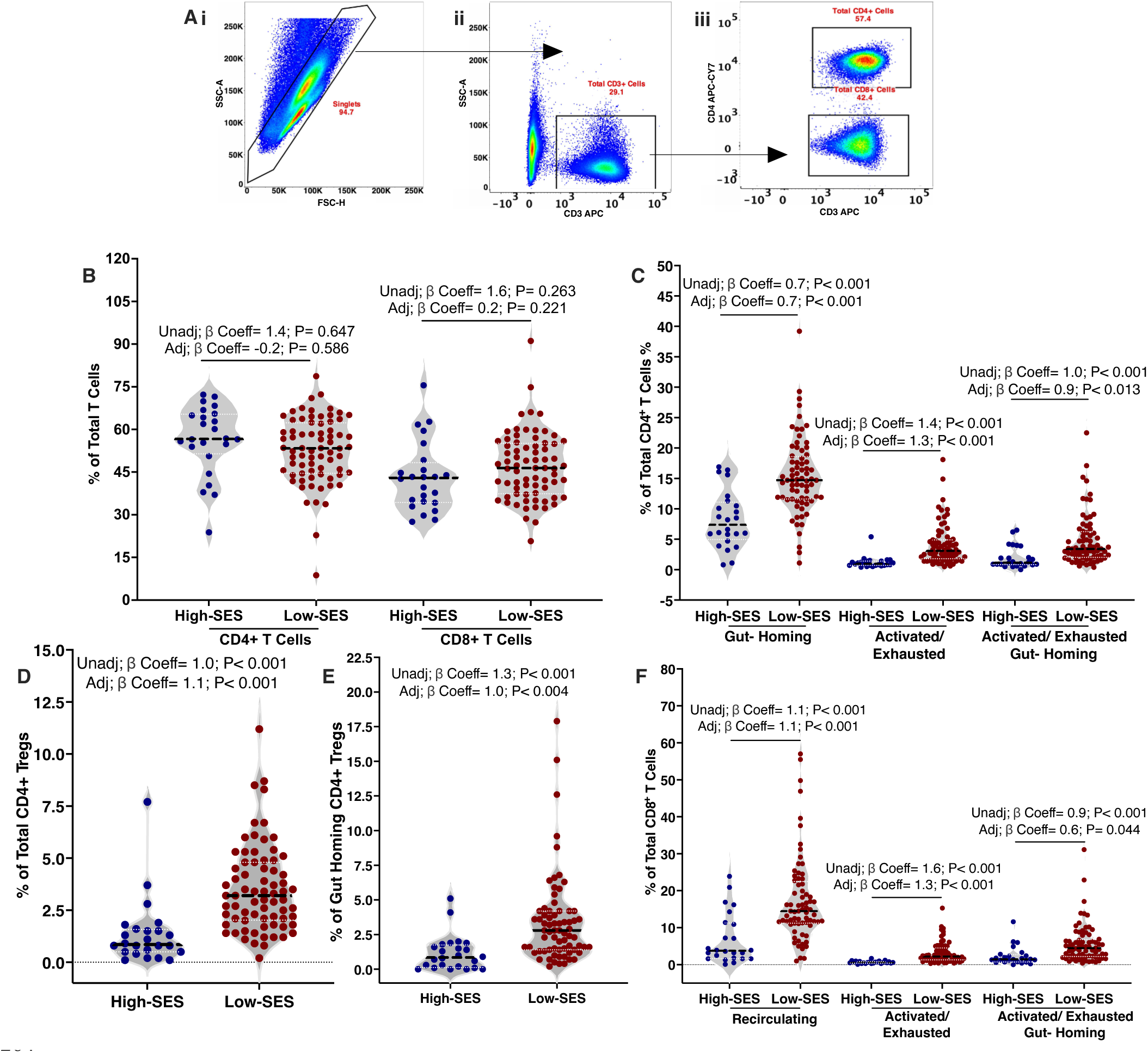
Adults from a Low-SES community have higher percentages of gut-homing and exhausted CD4^+^ and CD8^+^ T cells and Treg than those from the High-SES group. A. Total CD4 and CD8+ T cell flow cytometry gating strategy. Showing steps used in the identification of CD4+ and CD8+ T cells as percentages (left-to-right) i. Singlets (FSC-A versus FSC-H). ii. Total CD3+ T cell gate (SSC-A versus CD3-APC). iii. Total CD4+ (CD3+CD4+) and CD8+ (CD3+CD4-) T cells; Each gate is presented as a percentage of the population of interest. B. Percentages of total CD4+ (CD3^+^CD4^+^) and total CD8+ T cells (CD3^+^CD4-) in the High- and Low-SES groups. C. Percentages of total gut-homing (α4β7^+^), activated/ exhausted (PD1^+^), and gut-homing activated/ exhausted (α4β7^+^PD1^+^) CD4+ T cells in the High- and Low-SES groups. D. Percentages of total CD4^+^ Tregs (CD127^-/low^FOXP3^+^) and gut-homing Tregs (α4β7^+^) in the High- and Low-SES groups. E. Percentages of total gut-homing, activated/ exhausted, and gut-homing activated/ exhausted CD8+ T cells in the High- and Low-SES groups; Gut-homing (α4β7^+^); Activated/ exhausted (PD-1^+^); Gut-homing activated/ exhausted; CD4^+^ Tregs (CD127^-or^ ^low^/ FOXP3^+^). Associations between T cell phenotype (percentages) and SES group were evaluated using both unadjusted and adjusted (age, sex, BMI, seasonality, alcohol consumption, and HIV status) fractional regression models with the High-SES group as the reference. BH-adjusted P values are shown, with values < 0.05 considered significant. High-SES: (n= 24) and Low-SES: (n= 72).

### Monocyte and neutrophil activation are inversely associated with duodenal morphology characteristic of severe EE

After identifying differences in individual monocyte, neutrophil, total T cell and Treg phenotypes between adults with EE from Low- and High-SES groups, we sought to identify shared patterns of these variables across both SES groups. Using Principal Components Analysis (PCA) including all monocyte, neutrophil and T cell phenotypes, we identified Principal Component (PC)1 (positively loaded with monocyte and neutrophil expression of CD86 and TLR-4), PC2 (monocyte expression of HLA-DR), PC3 (T cell activation/exhaustion) and PC4 (T cell gut homing), as the main PCs, accounting for 46.1, 15.9, 11.2 and 8.4 % of the variance observed, respectively; together accounting for 81.6 % of the variance in our immunophenotyping dataset (Fig. 6A). We went on to investigate whether the identified PCs were associated with duodenal morphometric assessments of EE severity (i.e. epithelial surface area, villus height, villus width, crypt depth). We found a significant negative linear association between PC1 and villus height (adj β= -5.7: 95% CI -8.6, -2.7; P< 0.001) and PC1 and epithelial surface area (adj β= -5.0: 95% CI -7.5, -2.5; p< 0.001) after adjusting for confounders (Fig. 6B and 6C; Table S3). There were significant positive linear associations between PC1 (adj β= 4.1: 95% CI 1.3, 6.9; p= 0.027), PC2 (adj β= 2.7: 95% CI 0.7, 4.6; p=0.029, and PC4 (adj β= 2.2: 95% CI 0.6, 3.8; p= 0.028) and villus width in the unadjusted analyses but these were not statistically significant after adjusting for confounders (Fig. 6D-F and Table. S3). No significant associations were found between PC3 and any of the duodenal morphometry characteristics of EE (Table S3).

**Figure 6:**
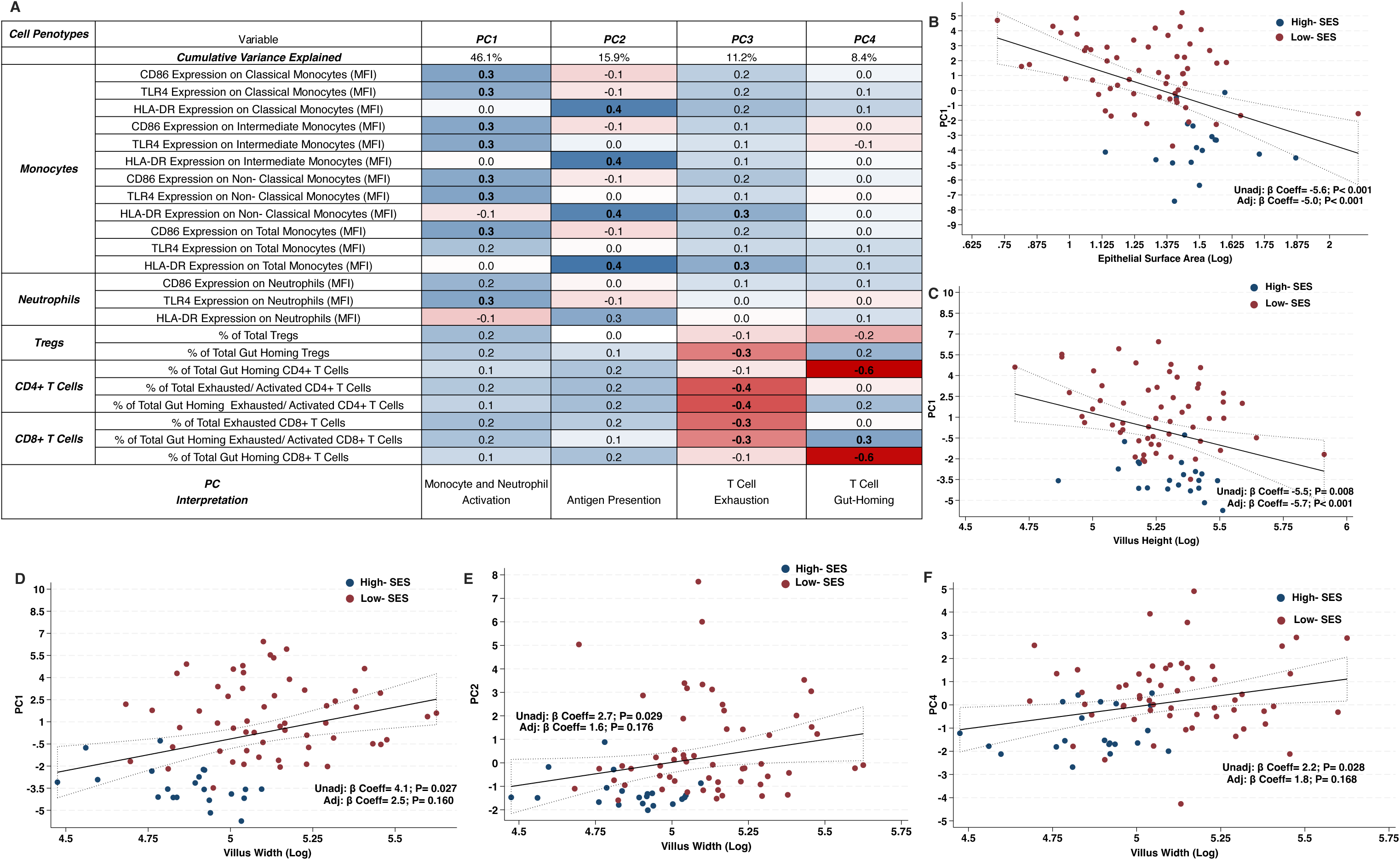
I**m**mune **cell phenotypes are associated with duodenal changes characteristic of EE severity:** A. The table shows PCs identified by PCA that included all innate and adaptive immune cell phenotyping variables across both the Low-SES (n=71) and High-SES (n=19) groups. Eigenvector loadings indicative of variables most strongly associated with the PC are colour coded; those with eigenvector loadings ≤ -3 are shaded dark blue and bolded; zero eigenvector loadings are shaded white; those with eigenvector loadings ≥ 3 are shaded dark red and bolded. Scatterplots are shown for PCs with statistically significant linear associations with duodenal morphology in unadjusted and/or adjusted (for age, sex, BMI, seasonality, alcohol consumption, and HIV status) linear regression models: B. PC1 with epithelial surface area; C PC1 with villus height; D. PC1 with villus width; E. PC2 with villus width; and F. PC4 with villus width. BH-adjusted P values are shown, with values < 0.05 considered significant. Solid lines indicate model fit and grey dotted boundaries denote the 95% CIs. Low-SES (n=57) and High-SES (n=17). % of Total CD4+ and CD8+ T cells were measured as a % of CD3+ T cells. % of Total Gut Homing, Total Activated/Exhausted and Total Gut-Homing Activated/Exhausted CD4+ and CD8+ T cells were measured as % of Total CD4 and CD8+ T cells, respectively; % of Total Tregs were measured as a % of Total CD4+ T cells; % of Total Gut-homing Tregs were measured as a % of Total Gut-homing CD4+ T cells.

## DISCUSSION

Inadequate sanitation is a hallmark of the residential areas where socio-economically disadvantaged people live, creating sharp contrasts in pathogen exposures within cities in Africa and Asia [34]. Previous studies have identified differences in duodenal characteristics and associated intestinal and systemic biomarkers of inflammation and microbial translocation between people in LMICs with EE and those without EE living in high-income countries [34]. Here, we show that socio-economic gradients translate into contrasts in gut health, microbial translocation, systemic inflammation, monocyte and neutrophil activation, T cell and Treg gut-homing, and activated/exhausted T cell phenotypes. We also show that monocyte and neutrophil activation (PC1) is associated with more severe EE. This suggests a potential bidirectional relationship in which systemic innate immune activation may contribute to EE severity and/or arise as a consequence of severe EE.

Gut health is characterised by long villi with shallow crypts, a large epithelial surface area and efficient digestion and nutrient absorption [35]. Villus blunting, defined by reduced villus height and increased villus width [1], is associated with accumulation of pro-inflammatory innate and adaptive immune cells in the lamina propria [13,36]. Increased crypt depth (crypt hyperplasia) may result from increased epithelial cell turnover, proliferation, and regeneration in response to inflammation [37]. In our study, villus width and crypt depth were higher in the Low-SES than in the High-SES group, confirming that these defining features of EE vary by SES.

In LMIC, plasma levels of systemic pro-inflammatory mediators, I-FABP, a biomarker of epithelial damage, and faecal levels of MPO, an enzyme produced by activated neutrophils, are elevated in children living in insanitary environments compared to those living in sanitary environments and are associated with linear growth [38–40]. Higher innate immune response-associated biomarkers of microbial translocation, sCD163 and sCD14 levels, have also been reported in adults with EE than in EE-free controls [23]. In our cohort, we found that I-FABP and MPO concentrations did not significantly differ between the Low-SES and the High-SES group after adjusting for relevant confounders. However, pro-inflammatory mediators, including plasma levels of CRP, LBP and sCD14, and EndoCAbs IgG and IgM, were higher in the Low-SES versus High-SES group, implicating innate and adaptive immune responses to microbial translocation [42]. Several studies indicate that EndoCAbs are associated with gram-negative microbial translocation [43,44], and previous work in Gambian children aged 8–48 weeks linked plasma EndoCab IgG to poorer growth, greater intestinal permeability and development of malnutrition at 18 months, suggesting possible EE-related malabsorption [45]. A suspected driver of EndoCAb responses in people with more severe EE is enteropathogen carriage; *Norovirus, Shigella, Campylobacter*, and enterotoxigenic *E. coli* which are correlated with greater intestinal damage and EE severity in young children in LMIC [46,47]. Together, these findings support a connection between SES, enteropathogen exposure, systemic immune activation, and EE severity.

Very few studies have investigated immune cell phenotypes in adults with EE. A key advance of our study is the characterisation of circulating monocyte, neutrophil, and T cell phenotypes in adults with EE from the Low- and High-SES urban communities within the same LMIC. Percentages of classical and non-classical monocyte subsets within the total monocyte pool were similar in the two SES groups, but there were fewer intermediate monocytes in the Low-SES group. Because these subsets are often considered part of a developmental continuum [48,49], with distinct roles [50,51], lower percentages of intermediate monocytes in the Low-SES adults may indicate shifts in monocyte differentiation and/or trafficking. All 3 subsets of monocytes from adults living in Low-SES environments had higher CD86 and TLR4 expression but lower HLA-DR expression than those from the High-SES environments. This suggests relatively higher sensitivity (TLR4) to bacterial PAMP such as LPS, and greater co-stimulatory capacity for T cells (CD86), but a lower capacity for antigen presentation to CD4+ T cells via HLA-DR than the High-SES group. Thus, this phenotype is a feature of endotoxin tolerance, which is characterised by dysfunctional antigen presentation [52,53]. Typically, neutrophils do not constitutively express CD86 and HLA-DR but can upregulate these markers under inflammatory conditions, allowing neutrophils to act as antigen-presenting cells in vitro and in vivo [54–58]. We observed higher CD86 and TLR-4 expression on neutrophils from the Low-SES group.

We found that adaptive immune cell phenotypes also differed between people living in Low-versus High-SES environments, with higher percentages of T cells expressing the gut-homing integrin (α4β7+), suggesting more active trafficking between the small intestine and the blood. In HIV, increased intestinal epithelial damage has been reported alongside increased mucosal macrophage proportions and reduced circulating monocyte abundance, with higher monocyte expression of β7 in antiretroviral therapy–naïve patients, consistent with immune-cell trafficking to the gut in people with enteropathy [59]. More adults in the Low-SES group were living with HIV than in the High-SES group, but differences in gut homing T cell percentages remained significant after adjustment for HIV status.

Percentages of activated/exhausted T cells also differed between the two SES groups; expression of PD-1, a surface immune checkpoint receptor upregulated on activated effector T cells in chronic inflammation, was higher on T cells from the Low-SES group. Higher frequencies of T cell effector memory, with increased expression of senescence-associated markers (CD57 and CD160), and upregulation of the immune-inhibitory checkpoint cytotoxic T-lymphocyte-associated protein 4 (CTLA-4) were reported in a Senegalese Low-SES group compared to the Netherlands control group [20]; but EE was not assessed in this cohort.

In addition to immune checkpoints, CD4^+^ Tregs (CD127^-orlow^/ FOXP3^+^) are essential regulators of chronic inflammation and immune cell activation. In this study, we found significantly greater abundance of total CD4^+^ Tregs (CD127^-/low^/ FOXP3^+^), and α4β7^+^ Tregs in the Low-versus High-SES group. Our data are consistent with findings from a murine model of EE (receiving both low protein/fat chow and CUMT8 *E. coli*), in which intestinal CD4^+^ Tregs were expanded and dampened effector T cell responses to oral vaccines [62]. People living with Crohn’s disease have significantly higher proportions of Tregs expressing GPR15, a colon-homing molecule, than Crohn’s-free controls [60] and there are higher circulating proportions of α4β7^+^ and GPR15^+^ Tregs in people with ulcerative colitis compared with both Crohn’s disease and healthy controls [61], suggesting that circulating gut-homing phenotypes vary across enteropathies. In ulcerative colitis, the suppressive function of CD4^+^CD25^+^CD127^low^FOXP3^+^ Tregs was lower in patients than in healthy controls, while the frequency, MFI of FOXP3, and Treg suppressive function were inversely correlated with disease activity [63]. It remains to be determined how the gut-homing Treg we identified in circulation relate to duodenal Treg reservoirs in EE or how their ability to control of intestinal inflammation might vary by SES.

There is growing evidence to support SES as a biological driver of altered systemic immune phenotypes [17–19]; but very few known studies have investigated EE severity as an underlying contributor to these differences. Using duodenal morphometry as a direct indicator of EE severity, we found that PC1, characterised by monocyte and neutrophil activation, was inversely associated with villus height and directly with epithelial surface area. Therefore, our findings suggest that circulating activated innate immune cell phenotypes may contribute to EE pathophysiology and/or reflect systemic responses to intestinal injury.

The findings of our study should be interpreted in light of several limitations. Our findings indicate that EE severity and immune cell phenotypes differ by SES, which encompasses multiple interrelated environmental exposures and health inequities that may influence gut health. Hence, although we adjusted for key confounders, several other SES-linked exposures may be driving these differences and remain to be identified. Given that EE is common in both High- and Low-SES communities in Zambia, including an EE-free comparator group from another geographic region would have added useful context but was beyond the scope of this study. Our immunophenotyping focused on a restricted set of phenotypic markers using a 6-parameter flow cytometer to avail of near-point-of-care laboratory facilities where our study was conducted; a more comprehensive phenotyping panel in future studies will allow for the delineation of more specific immune cell subsets in EE. We also used fixed cryopreserved buffy coat cells, precluding functional assays (e.g., Treg suppressive activity).

EE is widespread in LMICs; however, substantial SES disparities within these settings also exist. By directly quantifying duodenal morphometry alongside circulating biomarkers and immune cell phenotypes in clinically healthy adults in Lusaka, Zambia, we demonstrate that EE severity and its immunological imprint are not uniform within the same LMIC. Importantly, we show that activated monocytes and neutrophils linked to Low-SES are associated with histological features of EE severity, suggesting that systemic immune activation may contribute or may be a consequence of socioeconomic disparities in gut health of within LMIC.

## MATERIALS AND METHODS

### Study Participants and Study Design

This study is part of the Gastrointestinal (GI) Tools project, a clinical study that recruited healthy adults to undergo testing of gut functional capacity, as described elsewhere [65]. This is the cross-sectional immunology sub-study of GI Tools that included adults from Low-SES (n=76) and High-SES (n=26) communities in urban Lusaka, Zambia. Adults from the Low-SES group were recruited from Misisi compound, a known unplanned settlement locally referred to as a “shanty compound”. Most households in Misisi compound use pit latrines located very close to their water sources, whereas others share pit latrines with neighbouring households. They fetch water for household use from kiosks and store it in their homes. Adults from the High-SES group were enrolled from pre-selected residential areas (Rhodes Park, Longacres, Kabulonga, Ibex Hill, Chelston, Avondale, Sunningdale, Roma, Chamba Valley, Lilayi, New Kasama, etc.) through a Facebook advert and were only eligible if they had continuous access to clean, safe water and at least two flushable toilets per household. This study included only individuals aged 18 years or older who resided in the selected Low- or High-SES communities and provided at least one blood or stool sample for laboratory analyses. The study excluded women who were pregnant, individuals who had used antibiotics within the past 14 days, those experiencing diarrhoea at recruitment, and those with pre-existing gastrointestinal conditions or contraindications to endoscopy. Study nurses collected data on each participant’s demographics, existing clinical conditions and symptoms, household characteristics, occupation and relevant behaviours (e.g. smoking, alcohol consumption) at enrolment using study-specific questionnaires. SES characteristics were collected using a questionnaire that captured occupation, level of education, phone, house and car ownership and whether the household had electricity. Hygiene scores, a composite (total of ten) of overall household cleanliness, sanitation practices, water storage, food storage, and hand-washing practices and facilities, were calculated, with up to two points given for each item, as previously described [11]; A qualified study nurse, who had previously visited their homes, scored each participant (Low-SES). A score of 1–2 being Poor; 3–4 being Below acceptable; 5 being Acceptable / Average; 6–7 being Good; 8–9= Excellent and 10= Perfect. Participants’ weight and height were measured to calculate body mass index (BMI). Participants were enrolled from August 2022 through to February 2025; the season of recruitment (Rainy; December-April, Cool/ Dry; May-August, and Hot/ Dry; September to November), [66] was recorded for each participant.

### Sample Collection

Each participant provided a blood sample and a stool sample before the commencement of any other GI Tools study procedures. We collected 9mL of venous blood into Lithium heparin-treated collection tubes. Plasma was separated from buffy coat cells via centrifugation and stored at -80°C until analysis. The buffy coat cell layer containing immune cells was washed in phosphate-buffered saline (PBS), treated with Red Blood Cell lysis and fixation buffer (x1 BD FACS^TM^ lysing solution), washed and resuspended in freezing medium (10% dimethyl sulfoxide and 40% fetal bovine serum in PBS) and frozen at -80°C using a Mr. Frosty freezing container with isopropanol. Stool samples were collected in plain stool collection containers, aliquoted into cryovials, and stored at -80 °C until analysis. Within two days of blood and stool collection, participants underwent an endoscopic procedure. During this procedure, duodenal biopsies were collected using a Pentax Video Gastroscope (EG-2990i) and 2.8mm Single-Use Biopsy Standard Capacity Forceps (Boston Scientific). They were immediately oriented on cellulose acetate paper under a dissecting microscope and fixed in formalin for 24 hours. Thereafter, these biopsies were paraffin-embedded and stored at room temperature until sectioning.

### Duodenal Histology

Paraffin-embedded duodenal biopsies were sectioned (4μm) on a microtome (model CUT 4062; SLEE Medical GmbH, Mainz, Germany) and stained using Haematoxylin and Eosin (H&E). The slides were viewed using the Olympus VS120 slide-scanning microscope at 20x magnification. Measurements of the villus height, villus width, epithelial surface area (a two-dimensional representation of the epithelial perimeter along the brush border) and crypt depth were made in µm using the Virtual Slide Image Software (VSI), while epithelial surface area was expressed as a ratio to villus width. For each participant, measurements from multiple villi (2-8) were averaged for statistical analysis.

### Enzyme-linked immunosorbent Assay (ELISA)

Plasma immune biomarkers, sCD163, sCD14, LBP, AGP, CRP, and I-FABP (R&D Systems, Minneapolis, MN, USA), Endotoxin-core antibodies; IgG, IgA and IgM (Hycult Biotechnology B.V., Uden, The Netherlands), and faecal MPO (Immunodiagnostik AG, Bensheim, Germany) were measured using commercial ELISA kits (Table S4) following the manufacturer’s instructions. All ELISAs were analysed on a Biotek Synergy^TM^ LX Multi-Mode Microplate Reader.

### Flow Cytometry

Fixed cryopreserved buffy coat cell sample aliquots were separately stained using two fluorochrome-conjugated antibody panels. For monocyte and neutrophil activation analyses, we used surface staining with CD14-FITC, CD16-APC-Cy7, CD86-PerCP-Cy5.5, TLR4-PE, CD66b-APC, lymphocyte lineage (Lin; CD3, CD19, CD20, CD56)-APC (BioLegend, San Diego, CA, USA), and HLA-DR-PE-Cy7 (BD Biosciences, San Diego, CA, USA). For CD4+ Tregs and T cell analyses, we used CD3-APC, CD4-APC-Cy7, CD127-PE-Cy7 (BioLegend, San Diego, CA, USA), PD-1-PerCP Cy5.5 (BD Biosciences, San Diego, CA, USA), and α4β7-PE (BioLegend, San Diego, CA, USA) for surface marker staining, followed by permeabilisation (True-Nuclear™ Transcription Factor Buffer Set, BioLegend, San Diego, CA, USA) before intracellular staining for Foxp3-FITC (BioLegend, San Diego, CA, USA) (Table S3). For each sample, 300,000 events were acquired for the monocyte/neutrophil panel, and 400,000 events were acquired for the T cell panel on the Becton, Dickinson FACSVerse cytometer (488- and 633-nm lasers; 6 fluorochrome detection channels) using the FACSuite as the acquisition software (Table S4). All samples were analysed after the instrument performance quality control had passed. To account for voltage and spillover variations, single-stained compensation beads (UltraComp Beads, BD Biosciences, San Diego, CA, USA) and single-cell controls were used to apply compensation to all samples during analyses. Fluorescence Minus One controls (FMOs) were included in each batch to enable standardised hierarchical gating in FlowJo software version 10.8.1 (gating strategies are shown in Figs. S1, S2, and S3). Both panels were gated on single leukocytes. For the monocyte/neutrophil panel, monocyte subsets were identified within the total monocyte population (Lin^-^CD66b^-^HLA-DR^+^) based on CD14 and CD16 expression: classical monocytes (CD14^high^CD16^-^), intermediate monocytes (CD14^high^CD16^+^), and non-classical monocytes (CD14^low^CD16^+^). Neutrophils were identified as SSC-A^high^LinCD66b^+^CD16^+^. Median fluorescence intensities (MFIs) were used to quantify monocyte and neutrophil expression of TLR4, HLA-DR, and CD86. For the T cell panel, all CD3^+^CD4^-^ single cells (i.e. CD4-T cells) were assumed to be CD8^+^ T cells. The following phenotypes were identified: CD4^+^ Tregs (CD4^+^CD127^-/low^FOXP3^+^), CD4^+^ and CD8^+^ gut-homing T cells (α4β7^+^), CD4^+^ and CD8^+^ exhausted/ activated (PD-1^+^) T cells and CD4^+^ and CD8^+^ gut-homing exhausted/ activated (α4β7^+^PD-1^+^) T cells.

### Statistical Analysis

For demographic, clinical and household characteristics (Table 1), categorical variables were expressed as percentages and differences between the Low- and High-SES groups were evaluated using Fisher’s exact test. The Shapiro-Wilk test was employed to assess normality, and data were log-transformed prior to analyses as needed.

For immune biomarker (concentrations) and cell phenotypes (percentages and MFI) data, the Shapiro-Wilk test was employed to assess normality, and data were log-transformed as needed. To assess the magnitude of differences in concentrations and MFI between the two SES groups, we used linear regression with the High-SES group as the reference. Fractional regression was used to assess the magnitude of differences in cell percentages by SES, with High-SES as the reference group. Regression models were adjusted for potential confounder variables associated with immune changes, gut health and SES: participant age (continuous variable), sex (binary; male, female), HIV status (binary; positive, negative), alcohol consumption (binary; Yes, No), BMI (continuous variable), and season at time of sample collection (categorical; Rainy, Cool/Dry, Hot/Dry). Confounders were selected based on causal inference; i.e. the immune and gastrointestinal systems vary by age and sex, seasonal changes can alter disease transmission dynamics and pathogen infectivity [67], HIV status, BMI, and alcohol consumption differed by SES group and may affect intestinal permeability and differ by SES [68,69].

To reflect interactions among gut-homing immune cell phenotypes, we used Principal Component Analysis (PCA) to identify combinations of cell phenotypes that explain differences between the High- and Low-SES groups. All log-transformed monocyte, neutrophil, and T cell immunophenotyping data were standardised for both groups prior to entering the PCA. We selected the top 4 Principal Components (collectively explaining 76.3% of the total variance in our flow cytometry dataset across both SES groups); thereafter, the decreases or differences in each PC eigenvector loading were nearly constant. The immune cell phenotypes characterised by each PC were interpreted using eigenvector loadings, with variables loading onto the PC ≥ 0.3 and ≤ -0.3 considered meaningful contributors. We used linear regression to analyse associations between patterns of PC scores and duodenal morphometry (villus height, villus width, crypt depth, and epithelial surface area) as a measure of the association of immune cell phenotypes and EE severity. Models were adjusted for potential confounders as described above.

To account for multiple comparisons, p-values were adjusted using the Benjamini–Hochberg (BH) method. Statistical significance was defined as a BH-adjusted p-value < 0.05, and only adjusted p-values are presented. All statistical analyses were evaluated using Stata version 19. Figures were generated using Stata and GraphPad Prism version 10.

## Data Availability

All necessary information is included in the manuscript and supplementary material. Data can be obtained from the corresponding author, TNP, subject to required regulatory approvals and waivers. Since this data was generated as part of a PhD conducted under a project funded by a UKRI (MRC) grant, the data belong to the consortium, and additional approvals may be required.

## Ethics Statements

Ethical approvals were granted by the University of Zambia Biomedical Research Ethics Committee (UNZABREC) under reference numbers 2291-2021 and 4790-2023, and by the Zambian National Health Research Authority (NHRA) under reference numbers NHREB00001/02/02/2022 and NHREB002/01/04/2024, respectively. This study was conducted in accordance with the Declaration of Helsinki and Good Clinical and Laboratory Practices. All study participants provided written informed consent to participate.

## Acknowledgments

We would like to thank all study participants and their families for their participation in the study. We acknowledge intellectual input from all members of the GI Tools Consortium; all members are listed below.

GI Tools Consortium: Beatrice Amadi, Rosemary Banda, Ellen Besa, Claire D Bourke, Mutsa Bwakura-Dangarembizi, Ian Chisenga, Christine Edwards, Gary Frost, Isabel Garcia Perez, Mahek Jain, Leolin Katsidzira, Lydia Kazhila, Paul Kelly, Mirriam Kunaka, Kathryn Maitland, Nilanjan Mandal, Julian R Marchesi, Callum Melvin, Elena Monfort Sanchez, Douglas Morrison, Monica N Mweetwa, Mulima Mwiinga, Perpetual Ngalande, Tracy N Phiri, Joram M Posma, Ruari Robertson, Jose Ivan Serrano Contreras, Aaron Konzani Tembo, Alex J Thompson, James W Weatherill.

## Author contributions

Conceptualization: T.N.P, P.K, C.D.B

Methodology: T.NP, E.M, A.S, M.M, R.B, L.M, M.K, P.N, I.C, C.D.B, P.K

Investigation: T.N.P, E.M

Validation: T.N.P, E.M, C.D.B

Data curation: T.N.P, E.M, C.D.B

Visualization: T.N.P, E.M, C.D.B

Formal analyses: T.N.P, PK, C.D.B

Supervision: PK, C.D.B

Writing—original draft: T.N.P

Writing—review & editing: T.NP, E.M, A.S, L.M, M.K, P.N, I.C, C.D.B, P.K, GI TOOL team

