## SUPPLEMENTARY TABLES AND FIGURES for "Circulating Immune Cell Phenotypes are Associated with Socioeconomic Status and Severity of Environmental Enteropathy Among Zambian Adults"

### SUPPLEMENTARY MATERIAL

#### Supplementary Figures

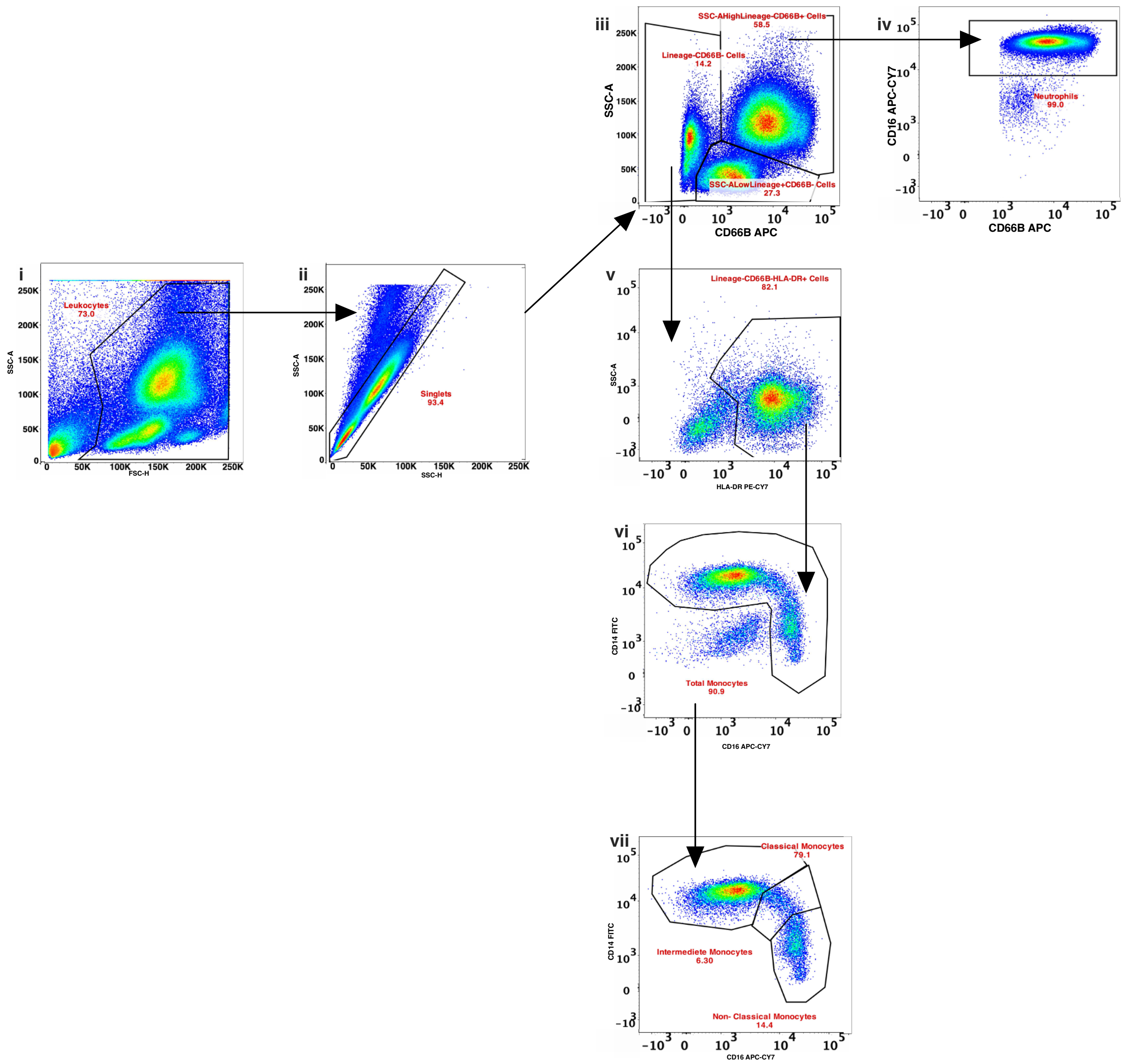


**Fig. S1. Monocyte and neutrophil gating strategy. Step-by-step flow of monocyte and neutrophil identification.** Using buffy coats, monocytes and neutrophils were identified by first gating on the Side scatter-Area (SSC-A) versus Forward scatter-Height (FSC-H) to get the Total leukocytes. This was followed by gating on the SSC-A versus SSC-H to get the Singlets. In the singlet gate, we identified Neutrophils using CD66B and CD16. Total monocytes were identified in the Lineage$^{-}$CD66B$^{-}$ gate using HLA-DR, and the monocyte subclasses within the total monocytes were identified by their distinct expression of CD14 and CD16. A Lineage (CD3, CD19, CD20, CD56) marker was used to exclude lymphocytes SSC-A$^{low}$Lineage$^{+}$CD66B$^{-}$ cells. The gating strategy shown is from one study participant; Numbers within the gates indicate the percentage of the cell population within the parent population (preceding gate) as an example. However, the total monocytes and total neutrophils are reported as a percentage of the singlet gate rather than the preceding gate. Numbers i-vii indicate the direction of the step-by-step gating.

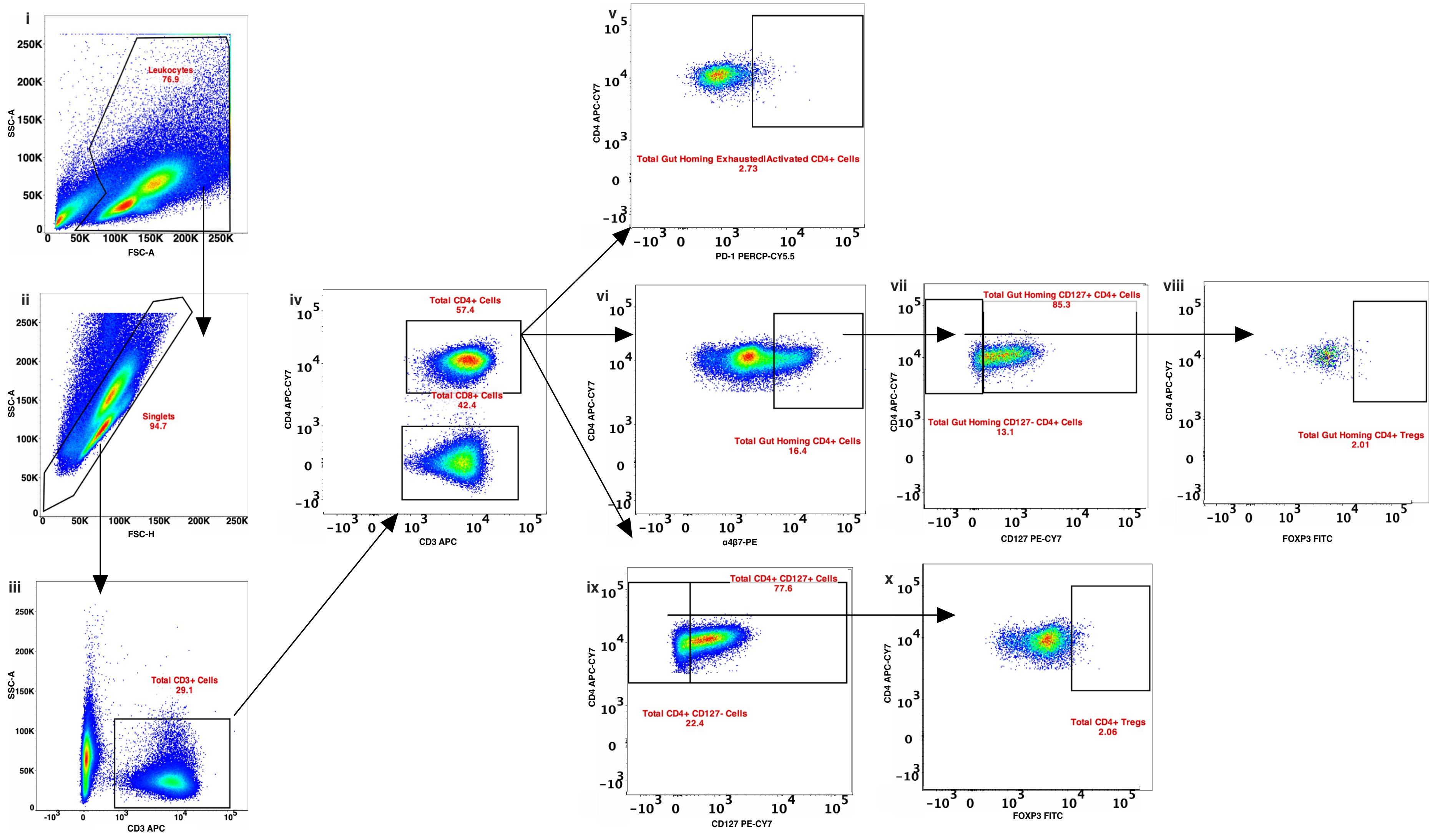

i
Leukocytes 76.9
SSC-A
FSC-A
ii
Singlets 94.7
SSC-A
FSC-H
iii
Total CD3+ Cells 29.1
SSC-A
CD3 APC
iv
Total CD4+ Cells 57.4
Total CD8+ Cells 42.4
CD4 APC-CY7
CD3 APC
v
Total Gut Homing Exhausted|Activated CD4+ Cells 2.73
CD4 APC-CY7
PD-1 PERCP-CY5.5
vi
Total Gut Homing CD4+ Cells 16.4
CD4 APC-CY7
α4β7-PE
vii
Total Gut Homing CD127+ CD4+ Cells 85.3
Total Gut Homing CD127- CD4+ Cells 13.1
CD4 APC-CY7
CD127 PE-CY7
viii
Total Gut Homing CD4+ Tregs 2.01
CD4 APC-CY7
FOXP3 FITC
ix
Total CD4+ CD127+ Cells 77.6
Total CD4+ CD127- Cells 22.4
CD4 APC-CY7
CD127 PE-CY7
x
Total CD4+ Tregs 2.06
CD4 APC-CY7
FOXP3 FITC

**Fig. S2. CD4+ T cell gating strategy.** Step-by-step flow of gut homing and activated/ exhausted $CD4^+$ T cell identification. Total leukocytes were identified using the SSC-A versus FSC-A. From the total leukocytes, we identified singlets using the FSC-A versus FSC-H. Using CD3+ as the marker, we identified T cells and later used CD4 to identify CD4+ T cells and all CD4- T cells were considered CD8+ T cells. In the CD4+ T cell population, we identified Total gut homing CD4+ T cells by their expression of $CD3^+CD4^+\alpha4\beta7^+$. Using $CD3^+CD4^+$PD1 expression, we identified activated/exhausted $CD4^+$ T cells. Using $CD3^+CD4^+\alpha4\beta7^+PD1^+$ expression, we Gut homing activated/exhausted CD4+ T cells. Further, using CD4+$CD127^{-/}$low FOXP3+ expression, we identified CD4+ Tregs; lastly, using $CD3^+CD4^+\alpha4\beta7^+CD127^{-/low}$ $FOXP3^+$ Expression, we identified gut-homing CD4+ Tregs within the Total CD4+ gut-homing population. The gating strategy shown is from one study participant; Numbers within gates indicate the percentage of the cell population within the parent population (preceding gate) as an example. The numbers indicate a sequential order of the gating. Numbers i-x indicate the direction of the step-by-step gating.

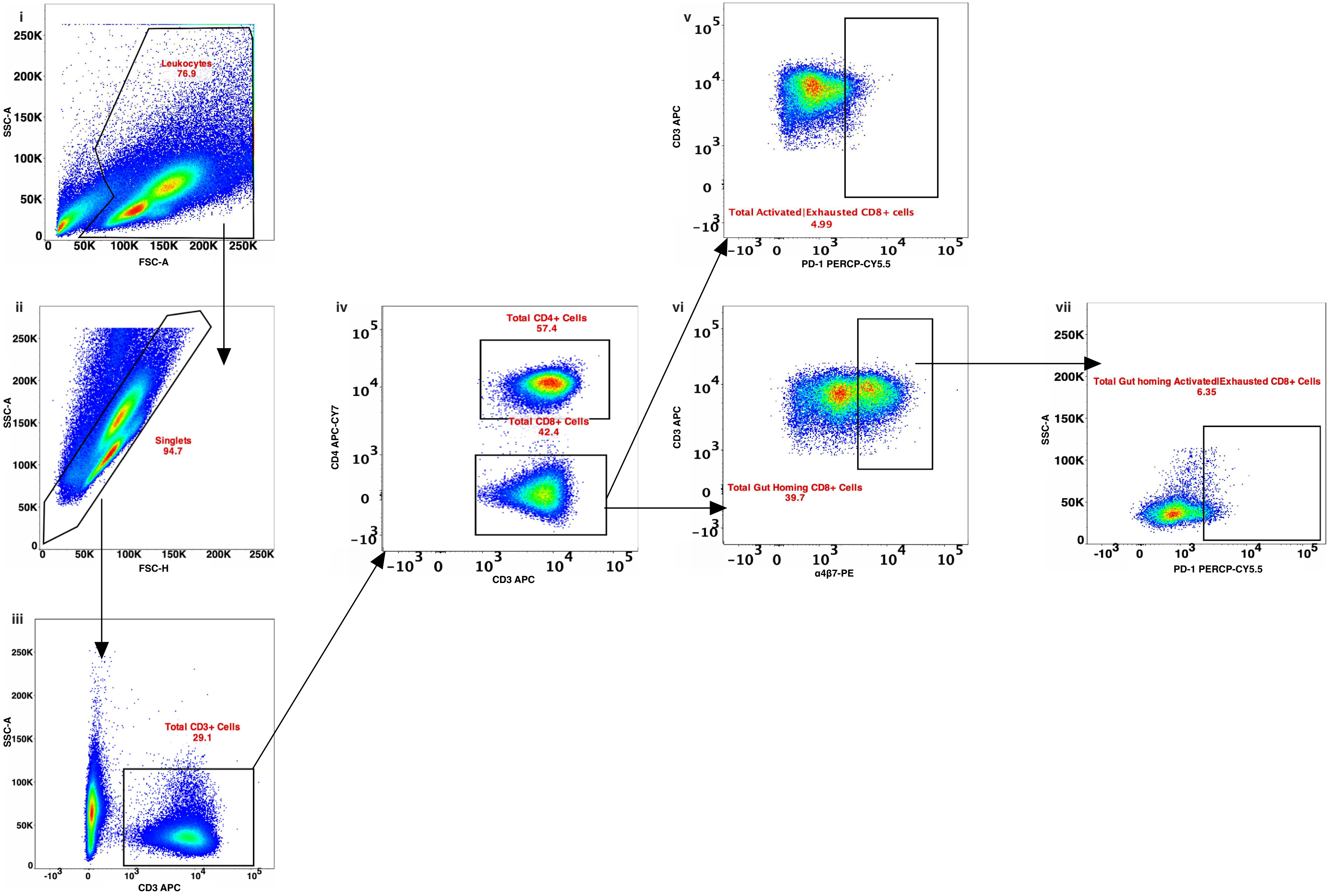

i
Leukocytes
76.9
SSC-A
FSC-A
ii
Singlets
94.7
SSC-A
FSC-H
iii
Total CD3+ Cells
29.1
SSC-A
CD3 APC
iv
Total CD4+ Cells
57.4
Total CD8+ Cells
42.4
CD4 APC-CY7
CD3 APC
v
Total Activated|Exhausted CD8+ cells
4.99
CD3 APC
PD-1 PERCP-CY5.5
vi
Total Gut Homing CD8+ Cells
39.7
CD3 APC
α4β7-PE
vii
Total Gut homing Activated|Exhausted CD8+ Cells
6.35
SSC-A
PD-1 PERCP-CY5.5

**Fig. S3. CD8+ T cell gating strategy.** Step-by-step flow of gut-homing and activated/ exhausted $CD8^+$ T cell identification. Total leukocytes were identified using the SSC-A versus FSC-A. From the total leukocytes, Singlets were identified using FSC-A versus FSC-H. In the singlets gate, CD3 was used to identify Total $CD3^+$ T cells. CD4 was used to identify CD4+ T cells; all cells that were CD4- negative were considered to be CD8+ T cells. iv. Using $CD3^+CD4^-\alpha4\beta7^+$ expression, we identified Total gut-homing CD8+ T cells. Using $CD3^+CD4^-PD1^+$ expression, we identified Total activated/exhausted $CD8^+$ T cells; lastly, using $CD3^+CD4^-\alpha4\beta7^+PD1^+$ expression, we identified total gut-homing activated/exhausted $CD8^+$ T cells. The gating strategy shown is from one study participant; Numbers within the gates indicate the percentage of the cell population within the parent population (preceding gate) as an example. The numbers indicate a sequential order of the gating. Numbers i-vii indicate the direction of the step-by-step gating.

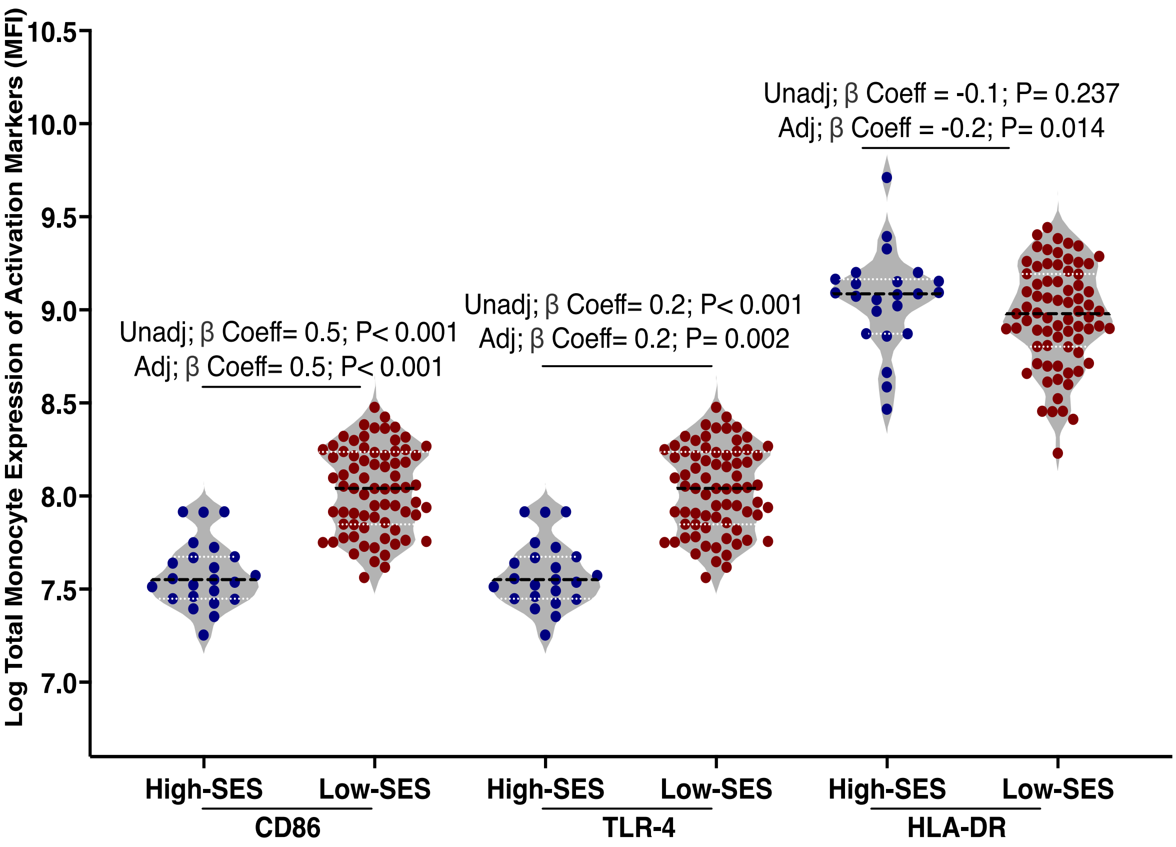


**Fig. S4. Total monocytes from adults from the low-SES group have lower expression of HLA-DR and higher expression of TLR4 and CD86 than those from the high-SES group.** CD86, TLR4 and HLA-DR expression on total monocytes (Lin-CD66b-HLA-DR+). Associations were assessed using both unadjusted and adjusted (for age, sex, BMI, seasonality, alcohol consumption, and HIV status) linear regression models with the high-SES group as the reference group. Benjamini-Hochberg adjusted p values are shown, with values < 0.05 considered significant. High-SES (n=23) and Low-SES (n=73).

### Supplementary Tables

| | *High-SES* | *Low-SES* | *P value* |
|---|---|---|---|
| ***Diarrhoea (past 14 days)*** | **n=26** | **n=75** | 1.000 |
| *Yes* | 0(0.0%) | 0(0.0%) | |
| *No* | 26(100%) | 75(100%) | |
| ***Disability*** | **n=26** | **n=75** | 1.000 |
| *Yes* | 0(0.0%) | 0(0.0%) | |
| *No* | 26(100.0%) | 75(100.0%) | |
| ***Past Caesarian Sections*** | **n=26** | **n=75** | 0.119 |
| *Yes* | 5(19.2%) | 5(6.7%) | |
| *No* | 21(80.8%) | 70(93.3%) | |
| ***Karnofsky performance status*** | **n=25** | **n=72** | 1.000 |
| *100* | 24(96.0%) | 67(93.1%) | |
| *90* | 1(4.0%) | 5(6.9%) | |
| ***Pallor*** | **n=24** | **n=73** | 1.000 |
| *Yes* | 0(0.0%) | 0(0.0%) | |
| *No* | 24(100.0%) | 73(100.0%) | |
| ***Jaundice*** | **n=24** | **n=73** | 1.000 |
| *Yes* | 0(0.0%) | 0(0.0%) | |
| *No* | 24(100.0%) | 73(100.0%) | |
| ***Cervical Lymphadenopathy*** | **n=24** | **n=75** | 1.000 |
| *Yes* | 0(0.0%) | 0(0.0%) | |
| *No* | 24(100.0%) | 75(100.0%) | |
| ***Goiter*** | **n=24** | **n=73** | 1.000 |
| *Yes* | 0(0.0%) | 1(1.4%) | |
| *No* | 24(100.0%) | 72(98.6%) | |
| ***Oral Candidiasis*** | **n=24** | **n=73** | 1.000 |
| *Yes* | 0(0.0%) | 0(0.0%) | |

| | | | |
|---|---|---|---|
| *No* | 24(100.0%) | 73(100.0%) | |
| ***Oral Kaposis Sarcoma*** | **n=24** | **n=73** | 1.000 |
| *Yes* | 0(0.0%) | 0(0.0%) | |
| *No* | 24(100.0%) | 73(100.0%) | |
| ***Eye Problem*** | **n=24** | **n=73** | 0.436 |
| *Yes* | 1(4.2%) | 1(1.4%) | |
| *No* | 23(95.8%) | 72(98.6%) | |
| ***BCG Scar*** | **n=24** | **n=73** | 0.010 |
| *Yes* | 21(87.5%) | 44(60.3%) | |
| *No* | 3(12.5%) | 29(39.7%) | |
| ***Pellagra Dermatosis*** | **n=24** | **n=75** | 1.000 |
| *Yes* | 0(0.0%) | 0(0.0%) | |
| *No* | 24(100.0%) | 75(100.0%) | |
| ***Hepatomegaly*** | **n=24** | **n=73** | 1.000 |
| *Yes* | 0(0.0%) | 0(0.0%) | |
| *No* | 24(100.0%) | 73(100.0%) | |
| ***Splenomegaly*** | **n=24** | **n=73** | 1.000 |
| *Yes* | 0(0.0%) | 0(0.0%) | |
| *No* | 24(100.0%) | 73(100.0%) | |
| ***Kidney Palpable*** | **n=24** | **n=73** | 1.000 |
| *Yes* | 0(0.0%) | 0(0.0%) | |
| *No* | 24(100.0%) | 73(100.0%) | |
| ***Pelvic Mass*** | **n=24** | **n=73** | 1.000 |
| *Yes* | 0(0.0%) | 0(0.0%) | |
| *No* | 24(100.0%) | 73(100.0%) | |

**Table S1. Clinical characteristics of study participants by group.** Comparisons between the low- and high-SES groups were made using Fisher's exact test; P values are shown, with values < 0.05 considered significant; Karnofsky performance status: 100: Normal, no complaints, no evidence of disease; and 90: Able to carry on normal activity, minor signs or symptoms of disease (69).

| *Parameter Measured* | | *Unadjusted* | | | *Adjusted* | |
|---|---|---|---|---|---|---|
| | *β-coeff* | *95%CI* | *P Value* | *β-coeff* | *95%CI* | *P Value* |
| ***Duodenal Morphometry*** | | | | | | |
| **Villus Height** | -0.1 | -0.2, 0.0 | 0.249 | -0.1 | -0.2, 0.0 | 0.050 |
| **Villus Width** | 0.3 | 0.2, 0.4 | <0.001 | 0.3 | 0.2, 0.4 | <0.001 |
| **Crypt Depth** | 0.2 | 0.1, 0.3 | <0.001 | 0.2 | 0.1, 0.3 | 0.005 |
| **Epithelial Surface Area** | -0.2 | -0.3, -0.1 | 0.003 | -0.3, | -0.4, -0.1 | 0.002 |
| ***Immune Biomarkers*** | | | | | | |
| **I-FABP[1]** | 0.3 | 0.1, 0.6 | 0.012 | 0. 2 | 0.0, 0.5 | 0.112 |
| **MPO[2]** | -0.2 | -0.6, 0.2 | 0.323 | -0.3 | -0.8, 0.2 | 0.194 |
| **CRP[1]** | 0.3 | -0.4, 1.1 | 0.346 | 0.9 | 0.2, 1.6 | 0.022 |
| **AGP[1]** | 0.1 | -0.1, 0.3 | 0.323 | 0.2 | 0.0, 0.3 | 0.112 |
| **sCD14[1]** | 1.6 | 1.4, 1.8 | <0.001 | 1.5 | 1.3, 1.7 | <0.001 |
| **sCD163[1]** | 0.2 | 0.0, 0.5 | 0.044 | 0.3 | 0.0, 0.5 | 0.056 |
| **LBP[1]** | 0.8 | 0.5, 1.0 | <0.001 | 0.7 | 0.4, 1.0 | <0.001 |
| **EndoCab IgG[1]** | 2.2 | 1.8, 2.6 | <0.001 | 2.2 | 1.7, 2.6 | <0.001 |
| **EndoCab IgA[1]** | 0.6 | 0.3, 0.9 | 0.002 | 0.4 | 0.0, 0.8 | 0.053 |
| **EndoCab IgM[1]** | 0.6 | 0.3, 0.9 | <0.001 | 0.5 | 0.2, 0.9 | 0.015 |
| ***Circulating Monocyte Phenotype*** | | | | | | |
| **% of Total Monocyte** | -0.1 | -0.3, 0.2 | 0.645 | -0.1 | -0.3, 0.2 | 0.786 |
| **% of Classical Monocytes** | -0.1 | -0.3, 0.2 | 0.565 | 0.0 | -0.3, 0.3 | 0.909 |

| | | | | | | |
|---|---|---|---|---|---|---|
| **% of Intermediate Monocytes** | -0.1 | -0.3, 0.1 | 0.386 | -0.2 | -0.4, 0.0 | 0.034 |
| **% of Non- Classical Monocytes** | 0.1 | -0.2, 0.4 | 0.504 | 0.0 | -0.3, 0.4 | 0.897 |
| **CD86 Expression on Total Monocytes** | 0.5 | 0.4, 0.6 | <0.001 | 0.5 | 0.4, 0.6 | <0.001 |
| **TLR4 Expression on Total Monocytes** | 0.2 | 0.1, 0.4 | <0.001 | 1.0 | 0.4, 1.5 | <0.001 |
| **HLADR Expression on Total Monocytes** | -0.1 | -0.2, 0.0 | 0.249 | -0.8 | -1.4, -0.3 | 0.007 |
| **CD86 Expression on Classical Monocytes** | 0.5 | 0.4, 0.6 | <0.001 | 0.5 | 0.4, 0.6 | <0.001 |
| **TLR4 Expression on Classical Monocytes** | 0.3 | 0.2, 0.3 | <0.001 | 0.3 | 0.2, 0.3 | <0.001 |
| **HLADR Expression on Classical Monocytes** | 0.0 | -0.2, 0.1 | 0.565 | -0.2 | -0.4, -0.1 | 0.011 |
| **CD86 Expression on Intermediate Monocytes** | 0.4 | 0.3, 0.5 | <0.001 | 0.5 | 0.4, 0.6 | <0.001 |
| **TLR4 Expression on Intermediate Monocytes** | 0.4 | 0.3, 0.4 | <0.001 | 0.4 | 0.3, 0.5 | <0.001 |
| **HLADR Expression on Intermediate Monocytes** | 0.0 | -0.2, 0.2 | 0.916 | 0.0 | -0.2, 0.2 | 0.897 |
| **CD86 Expression on Non- Classical Monocytes** | 0.4 | 0 .3, 0.5 | <0.001 | 0. 4 | 0.3, 0.5 | <0.001 |
| **TLR4 Expression on Non- Classical Monocytes** | 0.5 | 0.4, 0.6 | <0.001 | 0.5 | 0.4, 0.7 | <0.001 |
| **HLA-DR Expression on Non- Classical Monocytes** | -0.2 | -0.3, 0.0 | 0.066 | -0.2 | -0.3, 0.0 | 0.117 |
| ***Circulating Neutrophil Phenotype*** | | | | | | |
| **% of Neutrophil** | 0.0 | -0.3, 0.3 | 0.951 | 0.2 | -0.2, 0.5 | 0.466 |
| **TLR4 Expression on Neutrophils** | 0.5 | 0.3 0.6 | <0.001 | 0.5 | 0.4, 0.6 | <0.001 |
| **CD86 Expression on Neutrophils** | 0.3 | 0.2, 0.4 | <0.001 | 0.2 | 0.1, 0.3 | 0.002 |
| **HLA-DR Expression on Neutrophils** | -0.2 | -0.4, -0.1 | 0.009 | -0. 4 | -0.5, -0.2 | <0.001 |
| ***Circulating CD4+ T Cell Phenotype*** | | | | | | |
| **% of Total CD4 + T Cells** | -0.1 | -0.4, 0.2 | 0.645 | -0.1 | -0.5, 0.3 | 0.653 |

| | | | | | | |
|---|---|---|---|---|---|---|
| **% of Total Gut Homing CD4+ T Cells** | 0.7 | 0.4, 1.0 | <0.001 | 0.8 | 0.4, 1.1 | <0.001 |
| **% of Total Activated/Exhausted CD4+ T Cells** | 1.4 | 1.0, 1.9 | <0.001 | 1.1 | 0.7, 1.5 | <0.001 |
| **% of Total Gut Homing Activated/Exhausted CD4+ T Cells** | 1.0 | 0.5, 1.4 | <0.001 | 0.6 | 0.2, 1.1 | 0.012 |
| ***CD4+ Treg Phenotype*** | | | | | | |
| **% of Total Tregs** | 1.0 | 0.5, 1.5 | <0.001 | 1.1 | 0.6, 1.5 | <0.001 |
| **% of Total Gut Homing Tregs** | 1.3 | 0.7, 2.0 | <0.001 | 1.0 | 0.4, 1.6 | 0.004 |
| ***Circulating CD8+ T Cell Phenotype*** | | | | | | |
| **% of Total CD8 + T Cells** | 0.2 | -0.1, 0.5 | 0.297 | 0.2 | -0.1, 0.5 | 0.229 |
| **% of Total Gut Homing CD8+ T Cells** | 1.1 | 0.6, 1.6 | <0.001 | 1.1 | 0 .6, 1.6 | <0.001 |
| **% of Total Activated/Exhausted CD8+ T Cells** | 1.6 | 1.3, 1.9 | <0.001 | 1.3 | 0.9, 1.7 | <0.001 |
| **% of Total Gut Homing Activated/Exhausted CD8+ T Cells** | 0 .9 | 0.4, 1.4 | <0.001 | 0.6 | 0.1, 1.2 | 0.044 |

**Table S2. Differences in duodenal morphometry measurements, immune biomarkers, and immune cell phenotypes between the High- and Low-SES groups.** Associations were evaluated using both unadjusted and adjusted (for age, sex, BMI, seasonality, alcohol consumption, and HIV status) linear regression models with high-SES as the reference group. Benjamini-Hochberg adjusted P values are shown, with values < 0.05 considered significant. [1]Biomarkers measured in plasma. [2]Biomarker measured in stool. % of Total monocytes were measured as a % of singlets; % of Total neutrophils were measured as a % of singlets; % of Total CD4+ and CD8+ T cells were measured as a % of CD3+ T cells. % of Total Gut Homing, Total Activated/Exhausted and Total Gut-Homing Activated/Exhausted CD4+ and CD8+ T cells were measured as % of Total CD4 and CD8+ T cells, respectively; % of Total Tregs were measured as a % of Total CD4+ T cells; % of Total Gut-homing Tregs were measured as a % of Total Gut-homing CD4+ T cells.

| PC | Duodenal Morphometry | *Unadjusted* | | | *Adjusted* | | |
|---|---|---|---|---|---|---|---|
| | | ***β-coeff*** | ***95% CI*** | ***P value*** | ***β-coeff*** | ***95% CI*** | ***P value*** |
| **PC 1** | Log Villus Height | -5.5 | -8.8, -2.2 | 0.008 | -5.7 | -8.6, -2.7 | <0.001 |
| **PC2** | | -0.7 | -3.2, 1.8 | 0.705 | -1.6 | -3.9, 0.7 | 0.204 |
| **PC3** | | 0.3 | -1.7, 2.3 | 0.848 | 0.3 | -1.9, 2.4 | 0.854 |
| **PC4** | | -0.1 | -2.1, 1.9 | 0.931 | -0.6 | -2.7, 1.5 | 0.671 |
| **PC 1** | Log Villus Width | 4.1 | 1.3, 6.9 | 0.027 | 2.5 | -0.3, 5.2 | 0.160 |
| **PC2** | | 2.7 | 0.7, 4.6 | 0.029 | 1.6 | -0.4, 3.5 | 0.176 |
| **PC3** | | -1.1 | -2.7, 0.5 | 0.262 | -1.7 | -3.5, 0.0 | 0.149 |
| **PC4** | | 2.2 | 0.6 ,3.8 | 0.028 | 1.8 | 0.1, 3.6 | 0.168 |
| **PC 1** | Log Crypt Depth | 3.5 | 0.4, 6.5 | 0.075 | 2.3 | -0.8, 5.3 | 0.204 |
| **PC2** | | 1.9 | 0.1, 3.7 | 0.078 | 1.6 | -0.6, 3.7 | 0.204 |
| **PC3** | | -0.2 | -2.0, 1.5 | 0.848 | -0.2 | -2.2, 1.8 | 0.868 |
| **PC4** | | 2.3 | 0.1 ,4.4 | 0.082 | 1.7 | -0.3, 3.6 | 0.165 |
| **PC 1** | Log Epithelial Surface Area | -5.6 | -8.2, -2.9 | <0.001 | -5.0 | -7.5, -2.5 | <0.001 |
| **PC2** | | -1.4 | -3.4, 0.7 | 0.247 | -1.8 | -3.7, 0.1 | 0.144 |
| **PC3** | | 1.5 | -0.2, 3.1 | 0.122 | 1.7 | 0.0, 3.5 | 0.163 |
| **PC4** | | -1.6 | -3.2, 0.1 | 0.107 | -1.9 | -3.6, -0.2 | 0.165 |

**Table S3. Association of principal components (PCs) with duodenal morphometry measures.** Associations between PC scores and duodenal morphometry (villus height, villus width, crypt depth, and epithelial surface area) were assessed using both unadjusted and adjusted (for age, sex, BMI, seasonality, alcohol consumption, and HIV status) linear regression models with High-SES group as the reference. Benjamini-Hochberg adjusted P values are shown, with values < 0.05 considered significant. High-SES (n = 17) and low-SES (n = 57). Villus height (µm), Villus width (µm), crypt depth (µm), and epithelial surface area (ratio; epithelial surface area: villus height)

| | *Item Description* | *Catalogue Number* | *Supplier* |
|---|---|---|---|
| **ELISA kits** | | | |
| **MPO ELISA** | IDK® Myeloperoxidase (Stool, Urine) | K6630 | Immunodiagnostik |
| **I-FABP ELISA** | Human FABP2/I-FABP Quantikine ELISA Kit, | DFBP20 | Research and Development (R&D) Systems |
| **sCD163 ELISA** | Human CD163 Quantikine ELISA Kit | DC1630 | R&D Systems |
| **sCD14 ELISA** | Human CD14 DuoSet ELISA, | DY383-05 | R&D Systems |
| **LBP ELISA** | Human LBP Duoset ELISA, | DY870-05 | R&D Systems |
| **EndoCab IgG, IgM, IgA ELISA** | EndoCab®, Human, ELISA kit, | HK504-AGM | Hycult |
| **CRP ELISA** | Human C-Reactive Protein/CRP DuoSet ELISA, | DY1707 | R&D Systems |
| **AGP ELISA** | Human alpha 1-Acid Glycoprotein Quantikine ELISA Kit | DAGP00 | R&D Systems |
| **ELISA Ancillary Kit** | DuoSet ELISA Ancillary Reagent Kit 2 | DY008B | R&D Systems |
| **Flow cytometry reagents** | | | |
| **Anti-CD3 Antibody** | APC/Cyanine7 anti-human CD4/ SK3 | 344616 | Biolegend |
| **Anti-CD4 Antibody** | APC anti-human CD3/ Clone HIT3a | 300312 | Biolegend |
| **Anti-PD-1 Antibody** | BD Pharmingen™ PerCP-Cy™5.5 Mouse anti-Human CD279 (PD-1) Clone EH12.1 | 561273 | BD Biosciences |
| **Anti-α4β7 Antibody** | Human Integrin alpha 4 beta 7/LPAM-1 (Research Grade Vedolizumab Biosimilar) PE-conjugated Antibody | FAB10078P-100 | Biotechne |
| **Anti-FOXP3 Antibody** | FITC anti-human FOXP3; Clone 206D | 320106 | Biolegend |
| **Anti-CD127 Antibody** | PE/Cyanine7 anti-human CD127 (IL-7Rα); Clone A019D5 | 351320 | Biolegend |
| **Anti-HLA-DR Antibody** | Hu HLA-DR PE-Cy7 L243 (G46-6) 50Tst; PE CY7 | 560651 | BD Biosciences |
| **Anti-CD66b Antibody** | APC anti-human CD66b Antibody [Clone: G10F5] | 305118 | Biolegend |
| **Anti-CD16 Antibody** | APC/Cyanine7 anti-human CD16 Antibody [Clone: 3G8] | 302018 | Biolegend |
| **Anti-CD14 Antibody** | Anti-CD14 FITC, HCD14; FITC | 325604 | Biolegend |

| | | | |
|---|---|---|---|
| **Anti-lymphocyte Lineage Antibodies** | APC anti-human Lineage Cocktail (CD3, CD19, CD20, CD56) UCHT1; HIB19; 2H7; 5.1H11; APC | 363601 | Biolegend |
| **Anti-TLR4 Antibody** | Anti-CD284 (TLR4) (HTA125); PE | 312806 | Biolegend |
| **Anti-CD86 Antibody** | PerCP/Cyanine5.5 anti-human CD86 Antibody [Clone: BU63] | 374216 | Biolegend |
| **FBS** | Fetal Bovine Serum | F9665-100ML | SIGMA |
| **CS&T Beads** | BD FACSuite CS&T beads | 650622 | BD Sciences |
| **Compensation Beads** | UltraComp Beads | 01-2222-42 | BD Sciences |
| **DMSO** | Dimethyl sulfoxide | | |
| **PBS** | Phosphate Buffered Saline | 10010-015 | SIGMA |
| **FOXP3 Permeabilisation buffer** | True-Nuclear™ Transcription Factor Buffer Set | 424401 | Biolegend |
| **FOXP3 Staining buffer** | Cell staining buffer | 420201 | Biolegend |

**Table S4. Details of primary reagents and corresponding suppliers.**

| *Instruments and Equipment* | | | |
|---|---|---|---|
| **Instrument** | **Type/ Version** | **Software** | **Supplier** |
| **ELISA Plate Reader** | Biotek Reader | Gene5 software | Agilent |
| **Flow Cytometer** | BD FACSVERSE | FACSUITE | BD Sciences |
| **Gastroscope** | Video Gastroscope EG-2990i with EPK-i5000 Processor | N/A | Pentax Medical |
| **Biopsy Forceps** | Radial Jaw 4 Single-Use Biopsy Forceps 2.8mm Standard Capacity (M00513380) | N/A | Boston Scientific |
| **Microtome** | CUT 4062 Precision Manual Rotary Microtome | N/A | SLEE medical GmbH |
| **Imaging Microscope** | Olympus VS120 slide-scanning microscope | Virtual Slide Image (VSI) | Olympus |
| *Data Analysis Softwares* | | | |
| **Software** | **Version** | **Developer and supplier** | |
| **FlowJo** | Version 10 | FlowJo, Limited Liability Company (LLC) | |
| **Stata** | Version 17 | Stata Corp LLC | |
| **Graph Pad** | Version 10 | GraphPad Software | |

**Table S5. Details of equipment and software used for data collection and analysis.**
